# A multi-region discrete time chain binomial model for infectious disease transmission

**DOI:** 10.1101/2025.05.26.25328280

**Authors:** Pallab K. Sinha, Siuli Mukhopadhyay

## Abstract

Conventional mathematical models of infectious diseases often overlook the spatial spread of the disease, focusing only on local transmission. However, spatial propagation of various diseases has been observed mainly due to the movement of infectious individuals from one region to another. In this work, we propose a multiregion discrete-time chain binomial framework to model dependencies between infection time series from neighboring regions. The proposed model is based on the epidemiological principle that new infections arise from interactions between susceptible and previously infected individuals. To strengthen the statistical formulation, the probability of new infections within each spatial unit is modeled through a link function, by allowing it to depend on lagged local incidence, inter-regional transmission effects, and region-specific covariates. The framework also allows for higher-order temporal dependence through an AR(p) specification. To reduce complexity of the model as the number of regions increases, sparsity is imposed by using spatial contiguity matrices and k-nearest-neighbor structures. For estimating these multiregion models, an appropriate likelihood function maximization approach is proposed. A predictive likelihood based method is used to generate forecasts of disease counts in connected spatial units. In-depth analyses of two real measles datasets and simulation studies—incorporating seasonal patterns, different weight matrices across connected regions, and varying intervention strategies—are used to motivate the proposed modeling framework and demonstrate its ability to reproduce realistic outbreak dynamics. While the simulation studies demonstrate applicability of the general AR(p) approach, the real data analysis of measles shows that an AR(1) model is adequate to capture the temporal dependence.

## 1 Introduction

Spatio-temporal models of infectious diseases are essential to improve our understanding of epidemic dynamics as they are able to capture the simultaneous evolution of the disease over time and space. The spatial propagation of various infectious diseases has been documented multiple times throughout history in the epidemiological literature, for example, the 14th century plague in Europe, smallpox in the 1500s, measles in the United Kingdom in 1900s (Grenfell et al. [2001]), and most recently the Covid-19 pandemic (Kianfar et al. [2022]). This spatial propagation of infectious diseases can be attributed to the intermixture of infectious individuals from one region with healthy susceptibles from another region. In this work, our aim is to propose a novel spatio-temporal epidemiological model that can accurately predict and forecast counts of an infectious disease, in a multi-region setup. We integrate traditional epidemiological model, the chain binomial framework, with statistical techniques to develop a new approach that offers a comprehensive view of the disease dynamics over space and time.

Epidemic chain models of infectious disease transmission, such as the discrete-time chain binomial model, have been widely used in epidemiological studies to understand transmission of various diseases such as measles in a school setting (En’ko [1989]), household chickenpox transmission (Abbey [1952]), and ebola, swine fever and foot and mouth diseases [Ferrari et al., 2005]. In these models (Bailey [1957], Becker [2017]), the binomial formulation arises from the assumption that each potential exposure during an epidemic is an independent Bernoulli trial, resulting in either infection or continued susceptibility. The Reed–Frost model, a well-known example of a chain binomial model, assumes that the number of new infections in time *t* depends on the number of infectious and susceptible individuals at the previous time point *t* − 1. The chain structure of these models makes them suitable for parameter estimation using maximum likelihood methods. Recently, chain binomial models have been utilized in the epidemic modeling of COVID-19, where [Lefévre et al., 2021] formulated a generalized Reed–Frost model that included two classes of infections (symptomatic vs asymptomatic) for testing and vaccination, However, prior to this work, epidemic chain binomial models have not been utilized to examine the spatial dynamics of infectious disease transmission.

To model the spatial transmission of infections caused by movement of infectious individuals from one region coming into effective contact with the susceptibles of another region, we propose a new multi-region chain binomial framework. This model is based on the epidemiological principle that new infections are generated from the corresponding susceptible populations and previous infection counts. To enhance the statistical rigor of the proposed approach, we assume that the probability of new infections for each spatial unit are modeled through an appropriately chosen link function to depend on the lagged local and inter-regional incidence and region-specific time-dependent covariate effects. Instead of an auto regressive order 1 (AR(1)) structure as in conventional chain models, we allow for AR(p) dependence (Fokianos and Kedem [2004], Sathish et al. [2025]). The AR(p) generalization allows us to capture complex temporal structure beyond first-order dependence by accounting for possible delayed and accumulated disease transmission effects over multiple time lags. To address the rapid growth in model complexity as the number of spatial units increases, we incorporate spatial contiguity matrices and k-nearest-neighbor structures to impose sparsity on inter-regional transmission effects.

In-depth simulations are conducted to study the effect of disease transmission rates, number of spatial regions, spatial distance metrics, covariates and vaccine coverages on the performance of the proposed method under various scenarios depicting real life disease epidemics. Robustness of the proposed approach to misspecifications in the AR order, distance based weight functions and percentage of suceptibles are investigated. Comparison of the proposed model’s performance with the HHH model (Held et al. [2005]) is recorded for different sizes of the susceptible population. We evaluated the model prediction and forecasting performances using root mean squared error, prediction bias and coverage probabilities. We also illustrate the performance of the proposed model using two real world data sets. The first dataset consists of city-level measles incidence data from the United Kingdom spanning the period 1944–1966. This dataset represents the pre-vaccination era and has been extensively analyzed in the epidemiological and statistical literature. Seasonal variation, the baby boom effect, and city-specific live birth rates are known to be important covariates influencing measles dynamics in this period. Accordingly, we assume 2% to 20% of the city-specific populations to represent the corresponding susceptible populations. To demonstrate the applicability of the proposed method in the post-vaccination era, we further analyze a more recent district-level measles dataset from an Indian state covering the period 2014–2020. Due to the effect of vaccination campaigns, the second data set allows us to adjust the susceptible population of each district by accounting for vaccinated cohorts over time. For each real data set, comparison and subsequent selection of models are carried out by using various criterion functions such as mean squared errors based on various error residuals, chi-squared statistic, deviance, and Akaike Information criterion (AIC). We also use the predictive likelihood method (Mukhopadhyay and Sathish [2019]) to simultaneously forecast disease counts in all spatial units involved for both data sets. Both short-term and long-term forecasting performance are examined, and the corresponding summary metrics are shown to be related to population density and public transport efficiency within the spatial units considered. Although both of our real-world datasets are based on measles epidemics, the proposed approach is applicable to a broad class of directly transmitted diseases in which infection results in either long-term immunity or death, thereby requiring consideration of only susceptible and infectious compartments. Furthermore, diseases that progress rapidly without long latent periods, and therefore do not need explicit modeling of host birth and death dynamics in the equations [Ferrari et al., 2005], can also be effectively modeled.

Various spatio-temporal models have been proposed in the literature to study the dynamics of disease epidemics. Building on the SIR model of [Kermack and McKendrick, 1927], Hethcote [2000], Sattenspiel and Dietz [1995], Zakary et al. [2017] proposed a multiregion version of the discrete SIR model that accounts for the spread of infectious diseases across multiple geographical regions, incorporating the movement of the population between them. A time-series adaptation of the SIR model, known as the TSIR model [Grenfell et al., 2001], has been used extensively to model the spatial spread of diseases. Bjørnstad et al. [2002] examined measles transmission in connected regions, modeling the force of infection as a function of resident infectious hosts, immigrant infections, and the transmission rate, while, Xia et al. [2004] integrated the TSIR model with a gravity-based spatial interaction framework to describe measles transmission between different communities. Another series of models, namely the endemicepidemic models (Held et al. [2005]) or the HHH models have been used to study infectious diseases over space and time. These models separate infection dynamics into two components: an endemic component handling the influx of new infections from external sources, and an epidemic component covering the contagious nature of the disease by assuming the expected number of transmissions to be a function of earlier infections in time. Meyer and Held [2017] studied this framework in a spatiotemporal setting, modeling case counts at specific locations and times as Poisson or negative binomial variables, with means determined by autoregressive effects, spatial transmission, and time- or unit-specific factors. Similarly, Bauer and Wakefield [2018] modeled counts using a negative binomial distribution and incorporated random endemic, neighborhood, and spatial CAR effects. More recently, Bracher and Held [2022] explored alternative weighting schemes—such as shifted Poisson, linear decay, and geometric weights—while Lu and Meyer [2023] introduced a zero-inflated version of the HHH model to address excess zeros.

Although both TSIR and HHH models have been used in the literature to study infectious disease dynamics over space and time, they differ in their treatment of transmission mechanisms, susceptible populations, and incorporation of covariate effects. Our objective here is to provide an alternative modeling approach that combines the chain-binomial transmission mechanism with temporal, spatial, and covariate effects. The proposed framework retains the epidemiological interpretation that new infections arise through interactions between infectious and susceptible individuals, while allowing statistical modeling of factors that influence disease transmission and spread across connected spatial regions. Apart from TSIR and HHH, individual level models (ILMs) have also been used to study the spatio-temporal dynamics of infectious diseases (Deardon et al. [2010], Mahsin et al. [2022]). In ILMs, an infection kernel that represents risk factors/covariates associated with both infected and susceptible individuals is used to model the probability of a susceptible individual getting infected at a particular time point. Parameter estimation is performed within a Bayesian framework using Markov chain Monte Carlo (MCMC) methods. However, the ILM approach differs from the population-level model considered here. Our main modelling objective is to predict and forecast disease incidences over space and time using aggregated surveillance data, and we use a computationally less intensive likelihood-based approach based on maximum likelihood estimation (MLE).

The remainder of the manuscript is structured as follows. Section 2 introduces the chain binomial model and provide details on the proposed multivariate chain binomial model, along with parameter estimation, model selection statistics and a forecasting algorithm. In Sections 3, 4 and 5, we give simulation results and analyse two real-world datasets, respectively, to illustrate the application of the proposed approach to model disease epidemics over space and time. Finally, Sections 6 and 7 provides closing remarks and links for data sets and R program, respectively.

## 2 The chain binomial model - single region

We start by introducing the conventional chain binomial model and show steps to add statistical rigour. Suppose, a time series of disease counts have been obtained from a single region. The number of infections at time (*t* + 1) is modelled as:

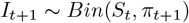

where *S*_*t*_ is the number of people at risk or susceptible to the disease at time *t*. The probability of infection *π*_*t*+1_ is generally chosen as a suitable nondecreasing function of the number of infections at time *t*, that is, *I*_*t*_, between 0 and 1. The susceptibles at time *t* + 1 is

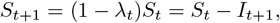

where *λ*_*t*_ is the force of infection and can be written as *λ*_*t*_ = *µI*_*t*_, *µ* is the effective rate of contact among infectious and susceptible individuals of that region. Thus, we obtain a generative equation 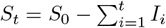, where *S*_0_ is the initial size of the susceptible population.

We deviate from a Markovian type of dependence as in [Ferrari et al., 2005], and assume that the probability of infections at time *t* may depend on infections until time lag *p* as in an AR(*p*) process. Also, we use a link function approach as in Fokianos and Kedem [2004] to incorporate the effect of long term disease trend, seasonal fluctuations, and time dependent covariates to model the probability of new infection.

Consider that the conditional distribution of *I*_*t*+1_|*H*_*t*_ is assumed to follow a 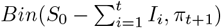 where *H*_*t*_ = *{***x**_*t*_, *S*_0_, *I*_*t*_, …, *I*_1_*}*. A logit link function is used as follows.

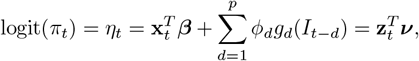

with 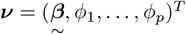, **z**_*t*_ = (**x**_*t*_, *I*_*t*−1_, …, *I*_*t*−*d*_), and *g*_*d*_(·) is any known function for each *d* (Fokianos and Kedem [2004]).

In the above equation, 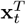 is assumed to be of the form *A*_*t*_ + *B*_*t*_ + *C*_*t*_, where *A*_*t*_ represents a possible long-term temporal trend component, *B*_*t*_ the seasonal components, and *C*_*t*_ may involve time dependent covariates such as vaccination coverages over time, live births, etc. The autoregressive part of the linear predictor is represented by the *p* terms, 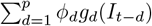. Similar forms of *η*_*t*_ have been used in the endemic-epidemic models as described in Held et al. [2005]. If required, lag terms corresponding to time dependent covariates may also be introduced. As an example, see lag terms of weather variables in the vector-borne disease model in Mukhopadhyay et al. [2019].

### 2.1 A new multi-region chain binomial model

To extend the conventional chain binomial model to a multi region model we make use of the multiple SIR type of framework as discussed in Zakary et al. [2017] for setting up the susceptibles. Suppose that there are *m* spatial regions and we observe disease counts 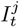, from the *j*th location, where *j* = 1, …, *m*. The susceptible population in the region *j* can then be written as 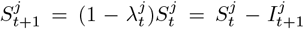, where 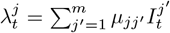 is the region-specific force of infection and 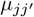 is the effective rate of contact between susceptible individuals in the region *j* with infectious individuals *j*^*′*^. Thus, the multi-region chain binomial model is as follows,

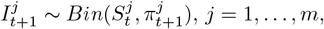

where the generative equation for the susceptible population *j*^*th*^ is updated to 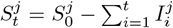, and 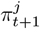 now represents the probability of infection at time (*t* + 1) in the region *j*.

As the *m* incidence counts observed are from neighboring spatial regions, they may be correlated with each other. Thus, we need to accommodate these dependencies over both time and space while modelling the probability of new infections in region *j*. We assume that the probability of new infection in region *j*, 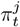, has AR(*p*) dependence not only on the disease counts of the same region *j*, i.e., 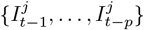, but also on the counts of the neighboring regions of *j*, that is, 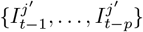, where *j*^*′*^ ∈ *N*_*j*_, *N*_*j*_ is a spatial neighborhood of *j*. Thus, the conditional distribution of 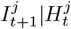 is as follows:

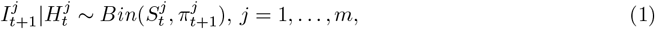

Where 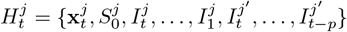, for all *j*^*′*^ ∈ *N*_*j*_.

Using a logit link function we model 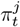 as

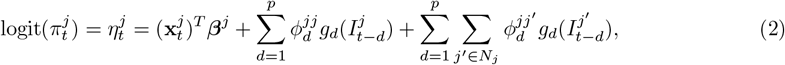

where 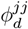 is the region-specific transmission parameter for region *j*, 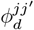 is the inter-region transmission parameter for region *j* and neighbouring region *j*^*′*^ and *g*_*d*_(·) as mentioned above is any known function for each *d* (Fokianos and Kedem [2004]). In our real data examples, we tried various forms of *g*(·) and the best fitting results were obtained for the logarithm function.

Here, we propose two choices to define *N*_*j*_.

1. Choice 1: *N*_*j*_ = {*j*^*′*^ : *j*^*′*^ ≠ *j*}, that is, assume that all surrounding regions may affect the counts of region *j* irrespective of sharing boundaries or being close to each other.
2. Choice 2: *N*_*j*_ = *{j*^*′*^ : *j*^*′*^ chosen using *k*-nearest neighbour rule*}*, for pre-specified values of *k*.

For Choice 1, note that even for 3 spatial regions and a AR(2) dependence, for each *j* we have 6 different 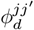 s, i.e. total 18 parameters. To reduce the number of inter region parameters, 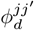, we assume that 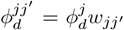, where 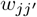 are distance-based weights. Thus, we use a *j*th region specific effect 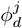 and weight it by its distance from its neighbour *j*^*′*^. We may model 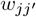 as a decreasing function of the distance between the centroids of the spatial regions *j* and *j*^*′*^, one popular choice is 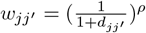, and *ρ* may be estimated from the data. For estimating the decay parameter, *ρ*, it is possible to use shifted poisson weights, linear decay or geometric weighting scheme as mentioned in Bracher and Held [2022]. For our computations, we also tried another set of weights 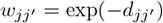. In simulation studies, we have studied the estimation results under various weight misspecifications. For the UK cities data set, we choose the weight function based on various goodness-of-fit statistics, see Table 9. Thus, using the distance-based weights the logit-AR(p) model is as follows for region *j*

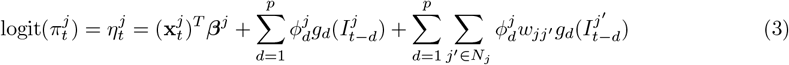

Specifically, 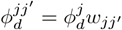, where for 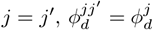 as 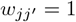.

### 2.2 Parameter Estimation

We use the maximum likelihood method for estimating the parameters, ***v*** = (***v***^1^, …, ***v***^*m*^),

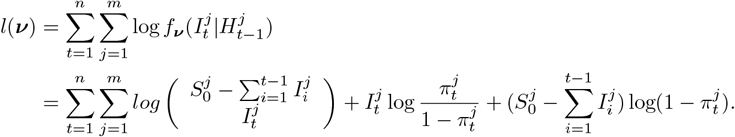

Using the logit link function, 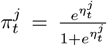, where 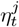 can be written as 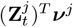. The Fisher scoring algorithm is used to find ***v*** iteratively as,

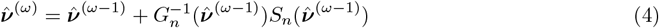

where *ω* denotes the *ω*^th^ iteration step in the estimation process.

The score function is,

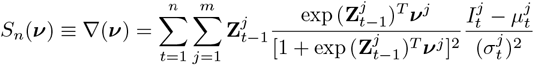

and the conditional covariance matrix *G*_*n*_(***v***) is given by:

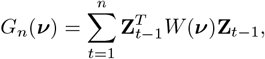

where *W* (***v***) is a diagonal matrix with elements 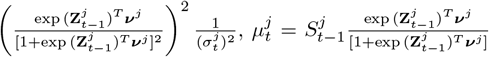 and 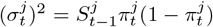. Fokianos and Kedem [1998] showed that 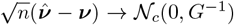 where *G* is the limiting term of *G*_*n*_ and *c* is the dimension of ***v***.

### 2.3 Susceptible population

The above MLE procedure depends on the values of the initial susceptible populations, 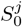. Determining a suitable initial susceptible population *S*_0_, that best describes the dynamics of the disease, is an important step to correctly analyze disease epidemics. In the epidemiological literature, it is generally assumed that *S*_0_ is the proportion not immune of the total population, i.e., *S*_0_ ≈ (1 − *p*)*N* where *p* denotes the immunity coverage and *N* denotes the population (Anderson and May [1991], Keeling and Rohani [2002]). Bjørnstad [2022] used from 2% to 20% of the total population as a candidate of the initial susceptible population. However, other methods such as estimating *S*_0_ using MLE as in Ferrari et al. [2005] have also been used. Very recently, Madden et al. [2024] used the TSIR model to get estimates of susceptibles in their neural network model setup. In this work, we follow the practices of the standard epidemic modeling literature and take 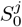, *j* = 1, …, *m* to be varying fractions of the region specific populations based on immunity or vaccination coverage. We investigate the effect of various values of 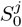 on the parameter estimates and coverage statistics in simulation studies. For the real data sets in later sections we present the results for 20% of the population densities, as is usually done for measles (Bjørnstad [2022], Ferrari et al. [2008]).

In vaccine preventable diseases (VPDs), particularly measles, the generative equation for 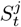 is further updated as

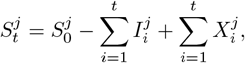

where 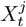 represents the non-immune birth cohort at time *t* and region *j* due to vaccination. For a vaccine preventable disease, if the number of scheduled doses, the efficacy of each dose, and coverages are known, the computation of 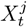 can be based on this information. Suppose, we have a VPD with one dose of vaccine, then the non-immune cohort is Chen et al. [2012]:

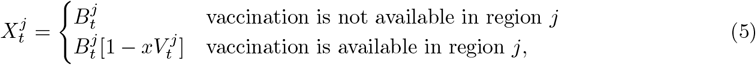

where 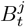 is the birth cohort, 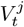 is the coverage of vaccination, and the efficacy of the vaccine is *x*.

### 2.4 Forecasting using a Predictive Likelihood

Once the model is selected, it is of interest to forecast future disease incidences and obtain intervals for these forecasts in the *m* regions. Based on past information up to the time point *n*, 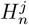, for all *j* regions, we estimate the density of the predicted value 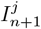 (one step ahead) using the predictive likelihood method (Mukhopadhyay and Sathish [2019]) as

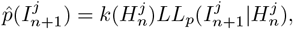

where *k*(*·*) is a normalizing constant and 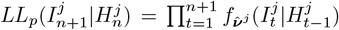. The forecast value of 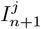 denoted by 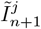, is arg max 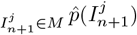, where 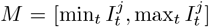. Note, 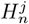 involves the counts of the *j*^th^ region as well as *j*^*′*^ *N*_*j*_. To generate, a set of *q* one-step ahead forecasts, the following steps are used:

Step 1: Generate the first forecast for all *j* regions; For *ω* = 1 and *j* = 1, …, *m* using the predictive likelihood,

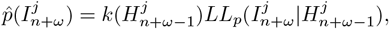

Step 2: To generate 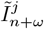, for *ω* = 2, …, *q* and *j* = 1, …, *m* we use the predictive likelihood

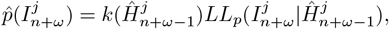

where, 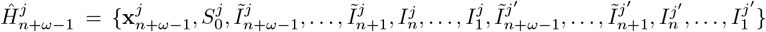 for all *j*^*′*^ ∈ *N*_*j*_.

To estimate the prediction interval for a set of one-step forward forecasts, we use the region of highest density of Hyndman [1995]. The (1 − *α*)100% HDR for 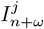, *ω* = 1, …, *q* is the region 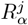, where 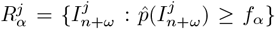, where *f*_*α*_ is the largest constant such that 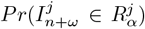 is at least (1 − *α*).

### 2.5 Model Evaluation

Various forms of the above multiregion chain binomial model were fitted to simulated and real data sets of disease counts. For evaluating these models, we used popular goodness-of-fit (GOF) statistics from the model selection literature like model deviance, chi-squared criterion, AIC, Bayesian Schwarz information criterion (BIC), various residuals (such as working, Pearson etc.) and mean square errors based on these residuals (Kedem and Fokianos [2005]).

Also, for evaluating model performance for in-sample predictions and forecasts and we used root mean square error, 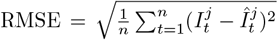, 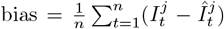, and mean absolute error, 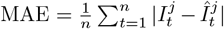 (Hyndman and Koehler [2006]). Forecasting coverage in the horizon *h* was also computed using Coverage 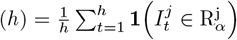, where **1**(*·*) is the indicator function and 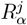 is the (1 − *α*)100% prediction interval from Section 2.4.

## 3 Simulation Studies

In this section, we present simulation results to illustrate the proposed approach. In the first subsection, data were simulated from equations 1 and 3. The primary objective was to investigate the effect of various forms of model misspecification on parameter estimation and predictive performance. The simulation results demonstrate that the proposed estimation procedure is robust to model misspecification.

To evaluate model performance, we examined the bias and variability of the parameter estimates, as well as the bias, MAE, coverage probability, and RMSE of the estimated counts for in-sample predictions and out-of-sample forecasts over two prediction horizons. We also compared the proposed model with the HHH model under two different sizes of the susceptible population. The results indicate that for a large susceptible pool, the proposed spatial chain binomial model performs comparably to the HHH model in terms of prediction and forecasting across different horizons. However, as the susceptible pool shrinks in size, the spatial chain binomial model consistently outperforms the HHH model, which relies on the Poisson assumption.

In the second subsection, data were generated from an multi region SIR (MSIR) model (Zakary et al. [2017]) under a vaccination scenario. The objective was to assess the ability of our proposed model approach to predict and forecast data arising from a fundamentally different data-generating mechanism. The second simulation study therefore provides information on the robustness and generalizability of the proposed model when the underlying epidemic process is different from the assumed model formulation.

### 3.1 Simulation Study 1

Data were generated using equations 1 and 3 for the following cases:

1. **Case 1, Large Susceptible pool:** *N*^*j*^ = 10^6^, 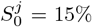 of *N*^*j*^ for *m* = 5; *p* = 2, *n* = 155 and 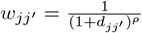,
2. **Case 2, Small Susceptible pool:** *N*^*j*^ = 10^4^, 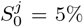 of *N*^*j*^ for *m* = 5; *p* = 2, *n* = 155 and 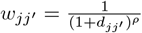,
3. **Case 3, Higher number of regions:** *N*^*j*^ = 10^6^, 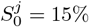 of *N*^*j*^ for *m* = 8; *p* = 2, *n* = 155 and 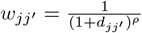,

In all the above cases, the time was assumed to be measured in biweeks and *ρ* was considered to be 2. Also trend and half yearly seasonal components were introduced in the model, specifically we assumed 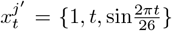, 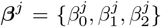, and *g*(*·*) to be logarithm in equations 1 and 3. We studied the performance of our proposed approach under the following misspecifications:

- AR order: AR(1) and AR(2)
- Weight function: 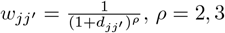 and 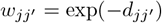
- 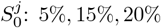 of *N*^*j*^

The simulations were repeated 100 times and the average performance was studied for these 100 simulations. For our proposed approach, we noted the average bias and average standard error (SE) of the model parameter estimates and the average bias, coverage probability, MAE, and RMSE values of the estimated counts for in-sample predictions. To assess forecasting capabilities, we forecasted for two horizons: one month or two biweeks (F1) and three months or six biweeks (F2) and again reported average MAE, bias, coverage and RMSE. To save space, the RMSE, MAE, bias and coverage values (both in-sample and forecasts) prediction of the incidence counts were averaged over the regions. The results for our proposed multi-region chain binomial model corresponding to cases 1,2 and 3 are shown in Tables 1, 2 and 3, respectively. We note from these tables that our proposed approach is robust across the various types of misspecifications listed above, with the highest log-likelihood values obtained for the true parameter model. Also, comparing the results of cases 1 and 3, we note that increasing *m* from 5 to 8 regions does not change our results much, except the coverage (for the true parameter case) for F2 in case 1 is 97% which decreases to 95% in case 3. Comparing results for cases 1 and 2, we note that the average bias in parameter estimates in higher for case 2 when compared to case 1.

**Table 1:**
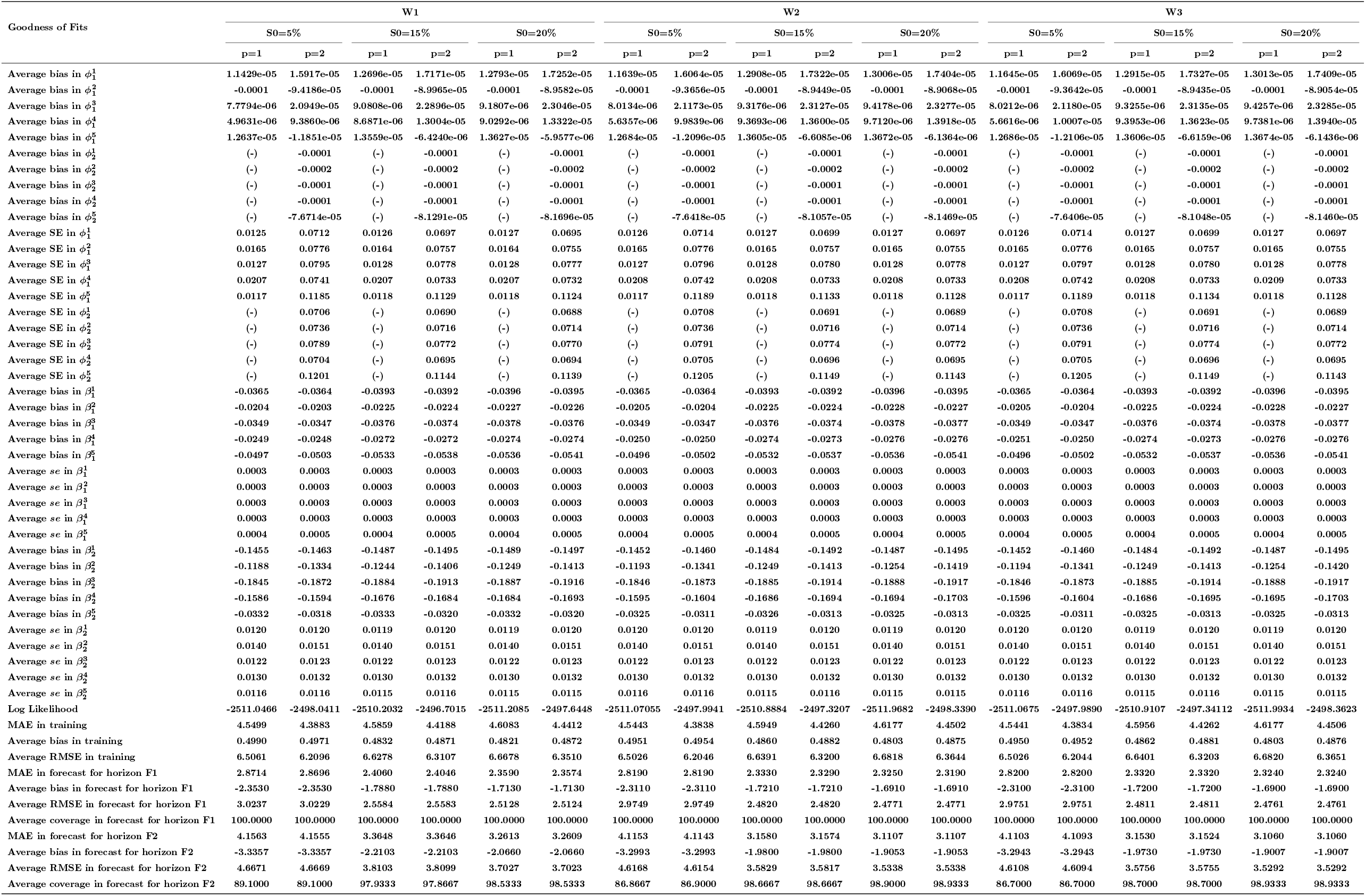
Parameter estimation and goodness of fit statistics of the proposed multi-region chain binomial model for case 1, Section 3.1.

| Goodness of Fits | W1 |  |  |  |  |  | W2 |  |  |  |  |  | W3 |  |  |  |  |  |
|---|---|---|---|---|---|---|---|---|---|---|---|---|---|---|---|---|---|---|
|  | S0=5% |  | S0=15% |  | S0=20% |  | S0=5% |  | S0=15% |  | S0=20% |  | S0=5% |  | S0=15% |  | S0=20% |  |
|  | p=1 | p=2 | p=1 | p=2 | p=1 | p=2 | p=1 | p=2 | p=1 | p=2 | p=1 | p=2 | p=1 | p=2 | p=1 | p=2 | p=1 | p=2 |
| Average bias in ϕ<sub>1</sub><sup>1</sup> | 1.1429e-05 | 1.5917e-05 | 1.2696e-05 | 1.7171e-05 | 1.2793e-05 | 1.7252e-05 | 1.1639e-05 | 1.6064e-05 | 1.2908e-05 | 1.7322e-05 | 1.3006e-05 | 1.7404e-05 | 1.1645e-05 | 1.6069e-05 | 1.2915e-05 | 1.7327e-05 | 1.3013e-05 | 1.7409e-05 |
| Average bias in ϕ<sub>1</sub><sup>2</sup> | -0.0001 | -9.4186e-05 | -0.0001 | -8.9965e-05 | -0.0001 | -8.9582e-05 | -0.0001 | -9.3656e-05 | -0.0001 | -8.9449e-05 | -0.0001 | -8.9068e-05 | -0.0001 | -9.3642e-05 | -0.0001 | -8.9435e-05 | -0.0001 | -8.9054e-05 |
| Average bias in ϕ<sub>1</sub><sup>3</sup> | 7.7794e-06 | 2.0949e-05 | 9.0808e-06 | 2.2896e-05 | 9.1807e-06 | 2.3046e-05 | 8.0134e-06 | 2.1173e-05 | 9.3176e-06 | 2.3127e-05 | 9.4178e-06 | 2.3277e-05 | 8.0212e-06 | 2.1180e-05 | 9.3255e-06 | 2.3135e-05 | 9.4257e-06 | 2.3285e-05 |
| Average bias in ϕ<sub>1</sub><sup>4</sup> | 4.9631e-06 | 9.3860e-06 | 8.6871e-06 | 1.3004e-05 | 9.0292e-06 | 1.3322e-05 | 5.6357e-06 | 9.9839e-06 | 9.3693e-06 | 1.3600e-05 | 9.7120e-06 | 1.3918e-05 | 5.6616e-06 | 1.0007e-05 | 9.3953e-06 | 1.3623e-05 | 9.7381e-06 | 1.3940e-05 |
| Average bias in ϕ<sub>1</sub><sup>5</sup> | 1.2637e-05 | -1.1851e-05 | 1.3559e-05 | -6.4240e-06 | 1.3627e-05 | -5.9577e-06 | 1.2684e-05 | -1.2096e-05 | 1.3605e-05 | -6.6085e-06 | 1.3672e-05 | -6.1364e-06 | 1.2686e-05 | -1.2106e-05 | 1.3606e-05 | -6.6159e-06 | 1.3674e-05 | -6.1436e-06 |
| Average bias in ϕ<sub>2</sub><sup>1</sup> | (-) | -0.0001 | (-) | -0.0001 | (-) | -0.0001 | (-) | -0.0001 | (-) | -0.0001 | (-) | -0.0001 | (-) | -0.0001 | (-) | -0.0001 | (-) | -0.0001 |
| Average bias in ϕ<sub>2</sub><sup>2</sup> | (-) | -0.0002 | (-) | -0.0002 | (-) | -0.0002 | (-) | -0.0002 | (-) | -0.0002 | (-) | -0.0002 | (-) | -0.0002 | (-) | -0.0002 | (-) | -0.0002 |
| Average bias in ϕ<sub>2</sub><sup>3</sup> | (-) | -0.0001 | (-) | -0.0001 | (-) | -0.0001 | (-) | -0.0001 | (-) | -0.0001 | (-) | -0.0001 | (-) | -0.0001 | (-) | -0.0001 | (-) | -0.0001 |
| Average bias in ϕ<sub>2</sub><sup>4</sup> | (-) | -0.0001 | (-) | -0.0001 | (-) | -0.0001 | (-) | -0.0001 | (-) | -0.0001 | (-) | -0.0001 | (-) | -0.0001 | (-) | -0.0001 | (-) | -0.0001 |
| Average bias in ϕ<sub>2</sub><sup>5</sup> | (-) | -7.6714e-05 | (-) | -8.1291e-05 | (-) | -8.1696e-05 | (-) | -7.6418e-05 | (-) | -8.1057e-05 | (-) | -8.1469e-05 | (-) | -7.6406e-05 | (-) | -8.1048e-05 | (-) | -8.1460e-05 |
| Average SE in ϕ<sub>1</sub><sup>1</sup> | 0.0125 | 0.0712 | 0.0126 | 0.0697 | 0.0127 | 0.0695 | 0.0126 | 0.0714 | 0.0127 | 0.0699 | 0.0127 | 0.0697 | 0.0126 | 0.0714 | 0.0127 | 0.0699 | 0.0127 | 0.0697 |
| Average SE in ϕ<sub>1</sub><sup>2</sup> | 0.0165 | 0.0776 | 0.0164 | 0.0757 | 0.0164 | 0.0755 | 0.0165 | 0.0776 | 0.0165 | 0.0757 | 0.0165 | 0.0755 | 0.0165 | 0.0776 | 0.0165 | 0.0757 | 0.0165 | 0.0755 |
| Average SE in ϕ<sub>1</sub><sup>3</sup> | 0.0127 | 0.0795 | 0.0128 | 0.0778 | 0.0128 | 0.0777 | 0.0127 | 0.0796 | 0.0128 | 0.0780 | 0.0128 | 0.0778 | 0.0127 | 0.0797 | 0.0128 | 0.0780 | 0.0128 | 0.0778 |
| Average SE in ϕ<sub>1</sub><sup>4</sup> | 0.0207 | 0.0741 | 0.0207 | 0.0733 | 0.0207 | 0.0732 | 0.0208 | 0.0742 | 0.0208 | 0.0733 | 0.0208 | 0.0733 | 0.0208 | 0.0742 | 0.0208 | 0.0733 | 0.0209 | 0.0733 |
| Average SE in ϕ<sub>1</sub><sup>5</sup> | 0.0117 | 0.1185 | 0.0118 | 0.1129 | 0.0118 | 0.1124 | 0.0117 | 0.1189 | 0.0118 | 0.1133 | 0.0118 | 0.1128 | 0.0117 | 0.1189 | 0.0118 | 0.1134 | 0.0118 | 0.1128 |
| Average SE in ϕ<sub>2</sub><sup>1</sup> | (-) | 0.0706 | (-) | 0.0690 | (-) | 0.0688 | (-) | 0.0708 | (-) | 0.0691 | (-) | 0.0689 | (-) | 0.0708 | (-) | 0.0691 | (-) | 0.0689 |
| Average SE in ϕ<sub>2</sub><sup>2</sup> | (-) | 0.0736 | (-) | 0.0716 | (-) | 0.0714 | (-) | 0.0736 | (-) | 0.0716 | (-) | 0.0714 | (-) | 0.0736 | (-) | 0.0716 | (-) | 0.0714 |
| Average SE in ϕ<sub>2</sub><sup>3</sup> | (-) | 0.0789 | (-) | 0.0772 | (-) | 0.0770 | (-) | 0.0791 | (-) | 0.0774 | (-) | 0.0772 | (-) | 0.0791 | (-) | 0.0774 | (-) | 0.0772 |
| Average SE in ϕ<sub>2</sub><sup>4</sup> | (-) | 0.0704 | (-) | 0.0695 | (-) | 0.0694 | (-) | 0.0705 | (-) | 0.0696 | (-) | 0.0695 | (-) | 0.0705 | (-) | 0.0696 | (-) | 0.0695 |
| Average SE in ϕ<sub>2</sub><sup>5</sup> | (-) | 0.1201 | (-) | 0.1144 | (-) | 0.1139 | (-) | 0.1205 | (-) | 0.1149 | (-) | 0.1143 | (-) | 0.1205 | (-) | 0.1149 | (-) | 0.1143 |
| Average bias in β<sub>1</sub><sup>1</sup> | -0.0365 | -0.0364 | -0.0393 | -0.0392 | -0.0396 | -0.0395 | -0.0365 | -0.0364 | -0.0393 | -0.0392 | -0.0396 | -0.0395 | -0.0365 | -0.0364 | -0.0393 | -0.0392 | -0.0396 | -0.0395 |
| Average bias in β<sub>1</sub><sup>2</sup> | -0.0204 | -0.0203 | -0.0225 | -0.0224 | -0.0227 | -0.0226 | -0.0205 | -0.0204 | -0.0225 | -0.0224 | -0.0228 | -0.0227 | -0.0205 | -0.0204 | -0.0225 | -0.0224 | -0.0228 | -0.0227 |
| Average bias in β<sub>1</sub><sup>3</sup> | -0.0349 | -0.0347 | -0.0376 | -0.0374 | -0.0378 | -0.0376 | -0.0349 | -0.0347 | -0.0376 | -0.0374 | -0.0378 | -0.0377 | -0.0349 | -0.0347 | -0.0376 | -0.0374 | -0.0378 | -0.0377 |
| Average bias in β<sub>1</sub><sup>4</sup> | -0.0249 | -0.0248 | -0.0272 | -0.0272 | -0.0274 | -0.0274 | -0.0250 | -0.0250 | -0.0274 | -0.0273 | -0.0276 | -0.0276 | -0.0251 | -0.0250 | -0.0274 | -0.0273 | -0.0276 | -0.0276 |
| Average bias in β<sub>1</sub><sup>5</sup> | -0.0497 | -0.0503 | -0.0533 | -0.0538 | -0.0536 | -0.0541 | -0.0496 | -0.0502 | -0.0532 | -0.0537 | -0.0536 | -0.0541 | -0.0496 | -0.0502 | -0.0532 | -0.0537 | -0.0536 | -0.0541 |
| Average *se* in β<sub>1</sub><sup>1</sup> | 0.0003 | 0.0003 | 0.0003 | 0.0003 | 0.0003 | 0.0003 | 0.0003 | 0.0003 | 0.0003 | 0.0003 | 0.0003 | 0.0003 | 0.0003 | 0.0003 | 0.0003 | 0.0003 | 0.0003 | 0.0003 |
| Average *se* in β<sub>1</sub><sup>2</sup> | 0.0003 | 0.0003 | 0.0003 | 0.0003 | 0.0003 | 0.0003 | 0.0003 | 0.0003 | 0.0003 | 0.0003 | 0.0003 | 0.0003 | 0.0003 | 0.0003 | 0.0003 | 0.0003 | 0.0003 | 0.0003 |
| Average *se* in β<sub>1</sub><sup>3</sup> | 0.0003 | 0.0003 | 0.0003 | 0.0003 | 0.0003 | 0.0003 | 0.0003 | 0.0003 | 0.0003 | 0.0003 | 0.0003 | 0.0003 | 0.0003 | 0.0003 | 0.0003 | 0.0003 | 0.0003 | 0.0003 |
| Average *se* in β<sub>1</sub><sup>4</sup> | 0.0003 | 0.0003 | 0.0003 | 0.0003 | 0.0003 | 0.0003 | 0.0003 | 0.0003 | 0.0003 | 0.0003 | 0.0003 | 0.0003 | 0.0003 | 0.0003 | 0.0003 | 0.0003 | 0.0003 | 0.0003 |
| Average *se* in β<sub>1</sub><sup>5</sup> | 0.0004 | 0.0005 | 0.0004 | 0.0005 | 0.0004 | 0.0005 | 0.0004 | 0.0005 | 0.0004 | 0.0005 | 0.0004 | 0.0005 | 0.0004 | 0.0005 | 0.0004 | 0.0005 | 0.0004 | 0.0005 |
| Average bias in β<sub>2</sub><sup>1</sup> | -0.1455 | -0.1463 | -0.1487 | -0.1495 | -0.1489 | -0.1497 | -0.1452 | -0.1460 | -0.1484 | -0.1492 | -0.1487 | -0.1495 | -0.1452 | -0.1460 | -0.1484 | -0.1492 | -0.1487 | -0.1495 |
| Average bias in β<sub>2</sub><sup>2</sup> | -0.1188 | -0.1334 | -0.1244 | -0.1406 | -0.1249 | -0.1413 | -0.1193 | -0.1341 | -0.1249 | -0.1413 | -0.1254 | -0.1419 | -0.1194 | -0.1341 | -0.1249 | -0.1413 | -0.1254 | -0.1420 |
| Average bias in β<sub>2</sub><sup>3</sup> | -0.1845 | -0.1872 | -0.1884 | -0.1913 | -0.1887 | -0.1916 | -0.1846 | -0.1873 | -0.1885 | -0.1914 | -0.1888 | -0.1917 | -0.1846 | -0.1873 | -0.1885 | -0.1914 | -0.1888 | -0.1917 |
| Average bias in β<sub>2</sub><sup>4</sup> | -0.1586 | -0.1594 | -0.1676 | -0.1684 | -0.1684 | -0.1693 | -0.1595 | -0.1604 | -0.1686 | -0.1694 | -0.1694 | -0.1703 | -0.1596 | -0.1604 | -0.1686 | -0.1695 | -0.1695 | -0.1703 |
| Average bias in β<sub>2</sub><sup>5</sup> | -0.0332 | -0.0318 | -0.0333 | -0.0320 | -0.0332 | -0.0320 | -0.0325 | -0.0311 | -0.0326 | -0.0313 | -0.0325 | -0.0313 | -0.0325 | -0.0311 | -0.0325 | -0.0313 | -0.0325 | -0.0313 |
| Average *se* in β<sub>2</sub><sup>1</sup> | 0.0120 | 0.0120 | 0.0119 | 0.0120 | 0.0119 | 0.0120 | 0.0120 | 0.0120 | 0.0119 | 0.0120 | 0.0119 | 0.0120 | 0.0120 | 0.0120 | 0.0119 | 0.0120 | 0.0119 | 0.0120 |
| Average *se* in β<sub>2</sub><sup>2</sup> | 0.0140 | 0.0151 | 0.0140 | 0.0151 | 0.0140 | 0.0151 | 0.0140 | 0.0151 | 0.0140 | 0.0151 | 0.0140 | 0.0151 | 0.0140 | 0.0151 | 0.0140 | 0.0151 | 0.0140 | 0.0151 |
| Average *se* in β<sub>2</sub><sup>3</sup> | 0.0122 | 0.0123 | 0.0122 | 0.0123 | 0.0122 | 0.0123 | 0.0122 | 0.0123 | 0.0122 | 0.0123 | 0.0122 | 0.0123 | 0.0122 | 0.0123 | 0.0122 | 0.0123 | 0.0122 | 0.0123 |
| Average *se* in β<sub>2</sub><sup>4</sup> | 0.0130 | 0.0132 | 0.0130 | 0.0132 | 0.0130 | 0.0132 | 0.0130 | 0.0132 | 0.0130 | 0.0132 | 0.0130 | 0.0132 | 0.0130 | 0.0132 | 0.0130 | 0.0132 | 0.0130 | 0.0132 |
| Average *se* in β<sub>2</sub><sup>5</sup> | 0.0116 | 0.0116 | 0.0115 | 0.0116 | 0.0115 | 0.0115 | 0.0116 | 0.0116 | 0.0115 | 0.0116 | 0.0115 | 0.0115 | 0.0116 | 0.0116 | 0.0115 | 0.0116 | 0.0115 | 0.0115 |
| Log Likelihood | -2511.0466 | -2498.0411 | -2510.2032 | -2496.7015 | -2511.2085 | -2497.6448 | -2511.07055 | -2497.9941 | -2510.8884 | -2497.3207 | -2511.9682 | -2498.3390 | -2511.0675 | -2497.9890 | -2510.9107 | -2497.34112 | -2511.9934 | -2498.3623 |
| MAE in training | 4.5499 | 4.3883 | 4.5859 | 4.4188 | 4.6083 | 4.4412 | 4.5443 | 4.3838 | 4.5949 | 4.4260 | 4.6177 | 4.4502 | 4.5441 | 4.3834 | 4.5956 | 4.4262 | 4.6177 | 4.4506 |
| Average bias in training | 0.4990 | 0.4971 | 0.4832 | 0.4871 | 0.4821 | 0.4872 | 0.4951 | 0.4954 | 0.4860 | 0.4882 | 0.4803 | 0.4875 | 0.4950 | 0.4952 | 0.4862 | 0.4881 | 0.4803 | 0.4876 |
| Average RMSE in training | 6.5061 | 6.2096 | 6.6278 | 6.3107 | 6.6678 | 6.3510 | 6.5026 | 6.2046 | 6.6391 | 6.3200 | 6.6818 | 6.3644 | 6.5026 | 6.2044 | 6.6401 | 6.3203 | 6.6820 | 6.3651 |
| MAE in forecast for horizon F1 | 2.8714 | 2.8696 | 2.4060 | 2.4046 | 2.3590 | 2.3574 | 2.8190 | 2.8190 | 2.3330 | 2.3290 | 2.3250 | 2.3190 | 2.8200 | 2.8200 | 2.3320 | 2.3320 | 2.3240 | 2.3240 |
| Average bias in forecast for horizon F1 | -2.3530 | -2.3530 | -1.7880 | -1.7880 | -1.7130 | -1.7130 | -2.3110 | -2.3110 | -1.7210 | -1.7210 | -1.6910 | -1.6910 | -2.3100 | -2.3100 | -1.7200 | -1.7200 | -1.6900 | -1.6900 |
| Average RMSE in forecast for horizon F1 | 3.0237 | 3.0229 | 2.5584 | 2.5583 | 2.5128 | 2.5124 | 2.9749 | 2.9749 | 2.4820 | 2.4820 | 2.4771 | 2.4771 | 2.9751 | 2.9751 | 2.4811 | 2.4811 | 2.4761 | 2.4761 |
| Average coverage in forecast for horizon F1 | 100.0000 | 100.0000 | 100.0000 | 100.0000 | 100.0000 | 100.0000 | 100.0000 | 100.0000 | 100.0000 | 100.0000 | 100.0000 | 100.0000 | 100.0000 | 100.0000 | 100.0000 | 100.0000 | 100.0000 | 100.0000 |
| MAE in forecast for horizon F2 | 4.1563 | 4.1555 | 3.3648 | 3.3646 | 3.2613 | 3.2609 | 4.1153 | 4.1143 | 3.1580 | 3.1574 | 3.1107 | 3.1107 | 4.1103 | 4.1093 | 3.1530 | 3.1524 | 3.1060 | 3.1060 |
| Average bias in forecast for horizon F2 | -3.3357 | -3.3357 | -2.2103 | -2.2103 | -2.0660 | -2.0660 | -3.2993 | -3.2993 | -1.9800 | -1.9800 | -1.9053 | -1.9053 | -3.2943 | -3.2943 | -1.9730 | -1.9730 | -1.9007 | -1.9007 |
| Average RMSE in forecast for horizon F2 | 4.6671 | 4.6669 | 3.8103 | 3.8099 | 3.7027 | 3.7023 | 4.6168 | 4.6154 | 3.5829 | 3.5817 | 3.5338 | 3.5338 | 4.6108 | 4.6094 | 3.5756 | 3.5755 | 3.5292 | 3.5292 |
| Average coverage in forecast for horizon F2 | 89.1000 | 89.1000 | 97.9333 | 97.8667 | 98.5333 | 98.5333 | 86.8667 | 86.9000 | 98.6667 | 98.6667 | 98.9000 | 98.9333 | 86.7000 | 86.7000 | 98.7000 | 98.7000 | 98.9333 | 98.9333 |

**Table 2:**
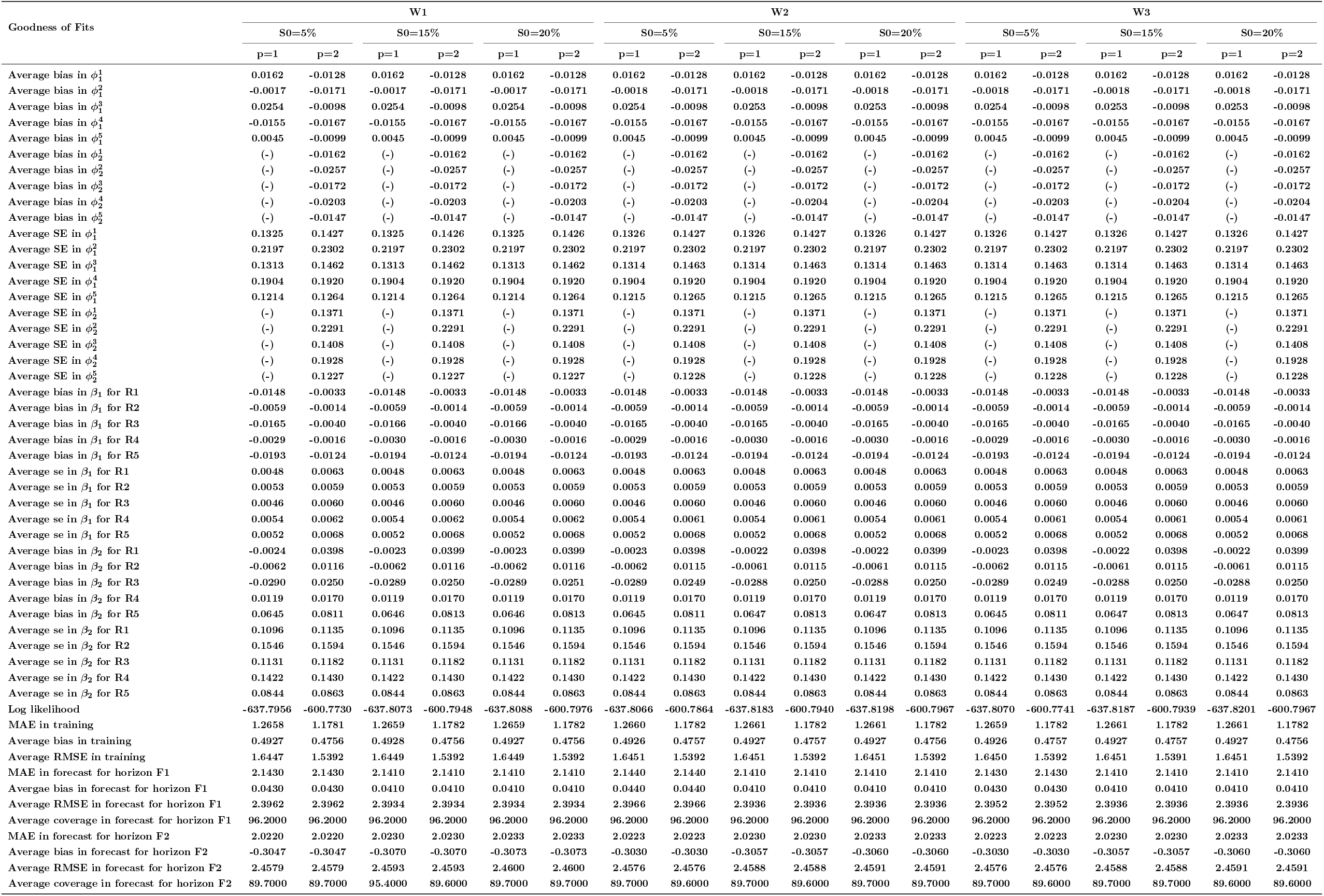
Parameter estimation and goodness of fit statistics of the proposed multi-region chain binomial model for case 2, Section 3.1.

| Goodness of Fits | W1 |  |  |  |  |  | W2 |  |  |  |  |  | W3 |  |  |  |  |  |
|---|---|---|---|---|---|---|---|---|---|---|---|---|---|---|---|---|---|---|
|  | S0=5% |  | S0=15% |  | S0=20% |  | S0=5% |  | S0=15% |  | S0=20% |  | S0=5% |  | S0=15% |  | S0=20% |  |
|  | p=1 | p=2 | p=1 | p=2 | p=1 | p=2 | p=1 | p=2 | p=1 | p=2 | p=1 | p=2 | p=1 | p=2 | p=1 | p=2 | p=1 | p=2 |
| Average bias in ϕ<sub>1</sub><sup>1</sup> | 0.0162 | -0.0128 | 0.0162 | -0.0128 | 0.0162 | -0.0128 | 0.0162 | -0.0128 | 0.0162 | -0.0128 | 0.0162 | -0.0128 | 0.0162 | -0.0128 | 0.0162 | -0.0128 | 0.0162 | -0.0128 |
| Average bias in ϕ<sub>1</sub><sup>2</sup> | -0.0017 | -0.0171 | -0.0017 | -0.0171 | -0.0017 | -0.0171 | -0.0018 | -0.0171 | -0.0018 | -0.0171 | -0.0018 | -0.0171 | -0.0018 | -0.0171 | -0.0018 | -0.0171 | -0.0018 | -0.0171 |
| Average bias in ϕ<sub>1</sub><sup>3</sup> | 0.0254 | -0.0098 | 0.0254 | -0.0098 | 0.0254 | -0.0098 | 0.0254 | -0.0098 | 0.0253 | -0.0098 | 0.0253 | -0.0098 | 0.0254 | -0.0098 | 0.0253 | -0.0098 | 0.0253 | -0.0098 |
| Average bias in ϕ<sub>1</sub><sup>4</sup> | -0.0155 | -0.0167 | -0.0155 | -0.0167 | -0.0155 | -0.0167 | -0.0155 | -0.0167 | -0.0155 | -0.0167 | -0.0155 | -0.0167 | -0.0155 | -0.0167 | -0.0155 | -0.0167 | -0.0155 | -0.0167 |
| Average bias in ϕ<sub>1</sub><sup>5</sup> | 0.0045 | -0.0099 | 0.0045 | -0.0099 | 0.0045 | -0.0099 | 0.0045 | -0.0099 | 0.0045 | -0.0099 | 0.0045 | -0.0099 | 0.0045 | -0.0099 | 0.0045 | -0.0099 | 0.0045 | -0.0099 |
| Average bias in ϕ<sub>2</sub><sup>1</sup> | (-) | -0.0162 | (-) | -0.0162 | (-) | -0.0162 | (-) | -0.0162 | (-) | -0.0162 | (-) | -0.0162 | (-) | -0.0162 | (-) | -0.0162 | (-) | -0.0162 |
| Average bias in ϕ<sub>2</sub><sup>2</sup> | (-) | -0.0257 | (-) | -0.0257 | (-) | -0.0257 | (-) | -0.0257 | (-) | -0.0257 | (-) | -0.0257 | (-) | -0.0257 | (-) | -0.0257 | (-) | -0.0257 |
| Average bias in ϕ<sub>2</sub><sup>3</sup> | (-) | -0.0172 | (-) | -0.0172 | (-) | -0.0172 | (-) | -0.0172 | (-) | -0.0172 | (-) | -0.0172 | (-) | -0.0172 | (-) | -0.0172 | (-) | -0.0172 |
| Average bias in ϕ<sub>2</sub><sup>4</sup> | (-) | -0.0203 | (-) | -0.0203 | (-) | -0.0203 | (-) | -0.0203 | (-) | -0.0204 | (-) | -0.0204 | (-) | -0.0203 | (-) | -0.0204 | (-) | -0.0204 |
| Average bias in ϕ<sub>2</sub><sup>5</sup> | (-) | -0.0147 | (-) | -0.0147 | (-) | -0.0147 | (-) | -0.0147 | (-) | -0.0147 | (-) | -0.0147 | (-) | -0.0147 | (-) | -0.0147 | (-) | -0.0147 |
| Average SE in ϕ<sub>1</sub><sup>1</sup> | 0.1325 | 0.1427 | 0.1325 | 0.1426 | 0.1325 | 0.1426 | 0.1326 | 0.1427 | 0.1326 | 0.1427 | 0.1326 | 0.1427 | 0.1326 | 0.1427 | 0.1326 | 0.1427 | 0.1326 | 0.1427 |
| Average SE in ϕ<sub>1</sub><sup>2</sup> | 0.2197 | 0.2302 | 0.2197 | 0.2302 | 0.2197 | 0.2302 | 0.2197 | 0.2302 | 0.2197 | 0.2302 | 0.2197 | 0.2302 | 0.2197 | 0.2302 | 0.2197 | 0.2302 | 0.2197 | 0.2302 |
| Average SE in ϕ<sub>1</sub><sup>3</sup> | 0.1313 | 0.1462 | 0.1313 | 0.1462 | 0.1313 | 0.1462 | 0.1314 | 0.1463 | 0.1314 | 0.1463 | 0.1314 | 0.1463 | 0.1314 | 0.1463 | 0.1314 | 0.1463 | 0.1314 | 0.1463 |
| Average SE in ϕ<sub>1</sub><sup>4</sup> | 0.1904 | 0.1920 | 0.1904 | 0.1920 | 0.1904 | 0.1920 | 0.1904 | 0.1920 | 0.1904 | 0.1920 | 0.1904 | 0.1920 | 0.1904 | 0.1920 | 0.1904 | 0.1920 | 0.1904 | 0.1920 |
| Average SE in ϕ<sub>1</sub><sup>5</sup> | 0.1214 | 0.1264 | 0.1214 | 0.1264 | 0.1214 | 0.1264 | 0.1215 | 0.1265 | 0.1215 | 0.1265 | 0.1215 | 0.1265 | 0.1215 | 0.1265 | 0.1215 | 0.1265 | 0.1215 | 0.1265 |
| Average SE in ϕ<sub>2</sub><sup>1</sup> | (-) | 0.1371 | (-) | 0.1371 | (-) | 0.1371 | (-) | 0.1371 | (-) | 0.1371 | (-) | 0.1371 | (-) | 0.1371 | (-) | 0.1371 | (-) | 0.1371 |
| Average SE in ϕ<sub>2</sub><sup>2</sup> | (-) | 0.2291 | (-) | 0.2291 | (-) | 0.2291 | (-) | 0.2291 | (-) | 0.2291 | (-) | 0.2291 | (-) | 0.2291 | (-) | 0.2291 | (-) | 0.2291 |
| Average SE in ϕ<sub>2</sub><sup>3</sup> | (-) | 0.1408 | (-) | 0.1408 | (-) | 0.1408 | (-) | 0.1408 | (-) | 0.1408 | (-) | 0.1408 | (-) | 0.1408 | (-) | 0.1408 | (-) | 0.1408 |
| Average SE in ϕ<sub>2</sub><sup>4</sup> | (-) | 0.1928 | (-) | 0.1928 | (-) | 0.1928 | (-) | 0.1928 | (-) | 0.1928 | (-) | 0.1928 | (-) | 0.1928 | (-) | 0.1928 | (-) | 0.1928 |
| Average SE in ϕ<sub>2</sub><sup>5</sup> | (-) | 0.1227 | (-) | 0.1227 | (-) | 0.1227 | (-) | 0.1228 | (-) | 0.1228 | (-) | 0.1228 | (-) | 0.1228 | (-) | 0.1228 | (-) | 0.1228 |
| Average bias in β<sub>1</sub> for R1 | -0.0148 | -0.0033 | -0.0148 | -0.0033 | -0.0148 | -0.0033 | -0.0148 | -0.0033 | -0.0148 | -0.0033 | -0.0148 | -0.0033 | -0.0148 | -0.0033 | -0.0148 | -0.0033 | -0.0148 | -0.0033 |
| Average bias in β<sub>1</sub> for R2 | -0.0059 | -0.0014 | -0.0059 | -0.0014 | -0.0059 | -0.0014 | -0.0059 | -0.0014 | -0.0059 | -0.0014 | -0.0059 | -0.0014 | -0.0059 | -0.0014 | -0.0059 | -0.0014 | -0.0059 | -0.0014 |
| Average bias in β<sub>1</sub> for R3 | -0.0165 | -0.0040 | -0.0166 | -0.0040 | -0.0166 | -0.0040 | -0.0165 | -0.0040 | -0.0165 | -0.0040 | -0.0165 | -0.0040 | -0.0165 | -0.0040 | -0.0165 | -0.0040 | -0.0165 | -0.0040 |
| Average bias in β<sub>1</sub> for R4 | -0.0029 | -0.0016 | -0.0030 | -0.0016 | -0.0030 | -0.0016 | -0.0029 | -0.0016 | -0.0030 | -0.0016 | -0.0030 | -0.0016 | -0.0029 | -0.0016 | -0.0030 | -0.0016 | -0.0030 | -0.0016 |
| Average bias in β<sub>1</sub> for R5 | -0.0193 | -0.0124 | -0.0194 | -0.0124 | -0.0194 | -0.0124 | -0.0193 | -0.0124 | -0.0194 | -0.0124 | -0.0194 | -0.0124 | -0.0193 | -0.0124 | -0.0194 | -0.0124 | -0.0194 | -0.0124 |
| Average se in β<sub>1</sub> for R1 | 0.0048 | 0.0063 | 0.0048 | 0.0063 | 0.0048 | 0.0063 | 0.0048 | 0.0063 | 0.0048 | 0.0063 | 0.0048 | 0.0063 | 0.0048 | 0.0063 | 0.0048 | 0.0063 | 0.0048 | 0.0063 |
| Average se in β<sub>1</sub> for R2 | 0.0053 | 0.0059 | 0.0053 | 0.0059 | 0.0053 | 0.0059 | 0.0053 | 0.0059 | 0.0053 | 0.0059 | 0.0053 | 0.0059 | 0.0053 | 0.0059 | 0.0053 | 0.0059 | 0.0053 | 0.0059 |
| Average se in β<sub>1</sub> for R3 | 0.0046 | 0.0060 | 0.0046 | 0.0060 | 0.0046 | 0.0060 | 0.0046 | 0.0060 | 0.0046 | 0.0060 | 0.0046 | 0.0060 | 0.0046 | 0.0060 | 0.0046 | 0.0060 | 0.0046 | 0.0060 |
| Average se in β<sub>1</sub> for R4 | 0.0054 | 0.0062 | 0.0054 | 0.0062 | 0.0054 | 0.0062 | 0.0054 | 0.0061 | 0.0054 | 0.0061 | 0.0054 | 0.0061 | 0.0054 | 0.0061 | 0.0054 | 0.0061 | 0.0054 | 0.0061 |
| Average se in β<sub>1</sub> for R5 | 0.0052 | 0.0068 | 0.0052 | 0.0068 | 0.0052 | 0.0068 | 0.0052 | 0.0068 | 0.0052 | 0.0068 | 0.0052 | 0.0068 | 0.0052 | 0.0068 | 0.0052 | 0.0068 | 0.0052 | 0.0068 |
| Average bias in β<sub>2</sub> for R1 | -0.0024 | 0.0398 | -0.0023 | 0.0399 | -0.0023 | 0.0399 | -0.0023 | 0.0398 | -0.0022 | 0.0398 | -0.0022 | 0.0399 | -0.0023 | 0.0398 | -0.0022 | 0.0398 | -0.0022 | 0.0399 |
| Average bias in β<sub>2</sub> for R2 | -0.0062 | 0.0116 | -0.0062 | 0.0116 | -0.0062 | 0.0116 | -0.0062 | 0.0115 | -0.0061 | 0.0115 | -0.0061 | 0.0115 | -0.0062 | 0.0115 | -0.0061 | 0.0115 | -0.0061 | 0.0115 |
| Average bias in β<sub>2</sub> for R3 | -0.0290 | 0.0250 | -0.0289 | 0.0250 | -0.0289 | 0.0251 | -0.0289 | 0.0249 | -0.0288 | 0.0250 | -0.0288 | 0.0250 | -0.0289 | 0.0249 | -0.0288 | 0.0250 | -0.0288 | 0.0250 |
| Average bias in β<sub>2</sub> for R4 | 0.0119 | 0.0170 | 0.0119 | 0.0170 | 0.0119 | 0.0170 | 0.0119 | 0.0170 | 0.0119 | 0.0170 | 0.0119 | 0.0170 | 0.0119 | 0.0170 | 0.0119 | 0.0170 | 0.0119 | 0.0170 |
| Average bias in β<sub>2</sub> for R5 | 0.0645 | 0.0811 | 0.0646 | 0.0813 | 0.0646 | 0.0813 | 0.0645 | 0.0811 | 0.0647 | 0.0813 | 0.0647 | 0.0813 | 0.0645 | 0.0811 | 0.0647 | 0.0813 | 0.0647 | 0.0813 |
| Average se in β<sub>2</sub> for R1 | 0.1096 | 0.1135 | 0.1096 | 0.1135 | 0.1096 | 0.1135 | 0.1096 | 0.1135 | 0.1096 | 0.1135 | 0.1096 | 0.1135 | 0.1096 | 0.1135 | 0.1096 | 0.1135 | 0.1096 | 0.1135 |
| Average se in β<sub>2</sub> for R2 | 0.1546 | 0.1594 | 0.1546 | 0.1594 | 0.1546 | 0.1594 | 0.1546 | 0.1594 | 0.1546 | 0.1594 | 0.1546 | 0.1594 | 0.1546 | 0.1594 | 0.1546 | 0.1594 | 0.1546 | 0.1594 |
| Average se in β<sub>2</sub> for R3 | 0.1131 | 0.1182 | 0.1131 | 0.1182 | 0.1131 | 0.1182 | 0.1131 | 0.1182 | 0.1131 | 0.1182 | 0.1131 | 0.1182 | 0.1131 | 0.1182 | 0.1131 | 0.1182 | 0.1131 | 0.1182 |
| Average se in β<sub>2</sub> for R4 | 0.1422 | 0.1430 | 0.1422 | 0.1430 | 0.1422 | 0.1430 | 0.1422 | 0.1430 | 0.1422 | 0.1430 | 0.1422 | 0.1430 | 0.1422 | 0.1430 | 0.1422 | 0.1430 | 0.1422 | 0.1430 |
| Average se in β<sub>2</sub> for R5 | 0.0844 | 0.0863 | 0.0844 | 0.0863 | 0.0844 | 0.0863 | 0.0844 | 0.0863 | 0.0844 | 0.0863 | 0.0844 | 0.0863 | 0.0844 | 0.0863 | 0.0844 | 0.0863 | 0.0844 | 0.0863 |
| Log likelihood | -637.7956 | -600.7730 | -637.8073 | -600.7948 | -637.8088 | -600.7976 | -637.8066 | -600.7864 | -637.8183 | -600.7940 | -637.8198 | -600.7967 | -637.8070 | -600.7741 | -637.8187 | -600.7939 | -637.8201 | -600.7967 |
| MAE in training | 1.2658 | 1.1781 | 1.2659 | 1.1782 | 1.2659 | 1.1782 | 1.2660 | 1.1782 | 1.2661 | 1.1782 | 1.2661 | 1.1782 | 1.2659 | 1.1782 | 1.2661 | 1.1782 | 1.2661 | 1.1782 |
| Average bias in training | 0.4927 | 0.4756 | 0.4928 | 0.4756 | 0.4927 | 0.4756 | 0.4926 | 0.4757 | 0.4927 | 0.4757 | 0.4927 | 0.4756 | 0.4926 | 0.4757 | 0.4927 | 0.4757 | 0.4927 | 0.4756 |
| Average RMSE in training | 1.6447 | 1.5392 | 1.6449 | 1.5392 | 1.6449 | 1.5392 | 1.6451 | 1.5392 | 1.6451 | 1.5392 | 1.6451 | 1.5392 | 1.6450 | 1.5392 | 1.6451 | 1.5391 | 1.6451 | 1.5392 |
| MAE in forecast for horizon F1 | 2.1430 | 2.1430 | 2.1410 | 2.1410 | 2.1410 | 2.1410 | 2.1440 | 2.1440 | 2.1410 | 2.1410 | 2.1410 | 2.1410 | 2.1430 | 2.1430 | 2.1410 | 2.1410 | 2.1410 | 2.1410 |
| Avergae bias in forecast for horizon F1 | 0.0430 | 0.0430 | 0.0410 | 0.0410 | 0.0410 | 0.0410 | 0.0440 | 0.0440 | 0.0410 | 0.0410 | 0.0410 | 0.0410 | 0.0430 | 0.0430 | 0.0410 | 0.0410 | 0.0410 | 0.0410 |
| Average RMSE in forecast for horizon F1 | 2.3962 | 2.3962 | 2.3934 | 2.3934 | 2.3934 | 2.3934 | 2.3966 | 2.3966 | 2.3936 | 2.3936 | 2.3936 | 2.3936 | 2.3952 | 2.3952 | 2.3936 | 2.3936 | 2.3936 | 2.3936 |
| Average coverage in forecast for horizon F1 | 96.2000 | 96.2000 | 96.2000 | 96.2000 | 96.2000 | 96.2000 | 96.2000 | 96.2000 | 96.2000 | 96.2000 | 96.2000 | 96.2000 | 96.2000 | 96.2000 | 96.2000 | 96.2000 | 96.2000 | 96.2000 |
| MAE in forecast for horizon F2 | 2.0220 | 2.0220 | 2.0230 | 2.0230 | 2.0233 | 2.0233 | 2.0223 | 2.0223 | 2.0230 | 2.0230 | 2.0233 | 2.0233 | 2.0223 | 2.0223 | 2.0230 | 2.0230 | 2.0233 | 2.0233 |
| Average bias in forecast for horizon F2 | -0.3047 | -0.3047 | -0.3070 | -0.3070 | -0.3073 | -0.3073 | -0.3030 | -0.3030 | -0.3057 | -0.3057 | -0.3060 | -0.3060 | -0.3030 | -0.3030 | -0.3057 | -0.3057 | -0.3060 | -0.3060 |
| Average RMSE in forecast for horizon F2 | 2.4579 | 2.4579 | 2.4593 | 2.4593 | 2.4600 | 2.4600 | 2.4576 | 2.4576 | 2.4588 | 2.4588 | 2.4591 | 2.4591 | 2.4576 | 2.4576 | 2.4588 | 2.4588 | 2.4591 | 2.4591 |
| Average coverage in forecast for horizon F2 | 89.7000 | 89.7000 | 95.4000 | 89.6000 | 89.7000 | 89.7000 | 89.7000 | 89.6000 | 89.7000 | 89.6000 | 89.7000 | 89.7000 | 89.7000 | 89.6000 | 89.7000 | 89.7000 | 89.6000 | 89.6000 |

**Table 3:**
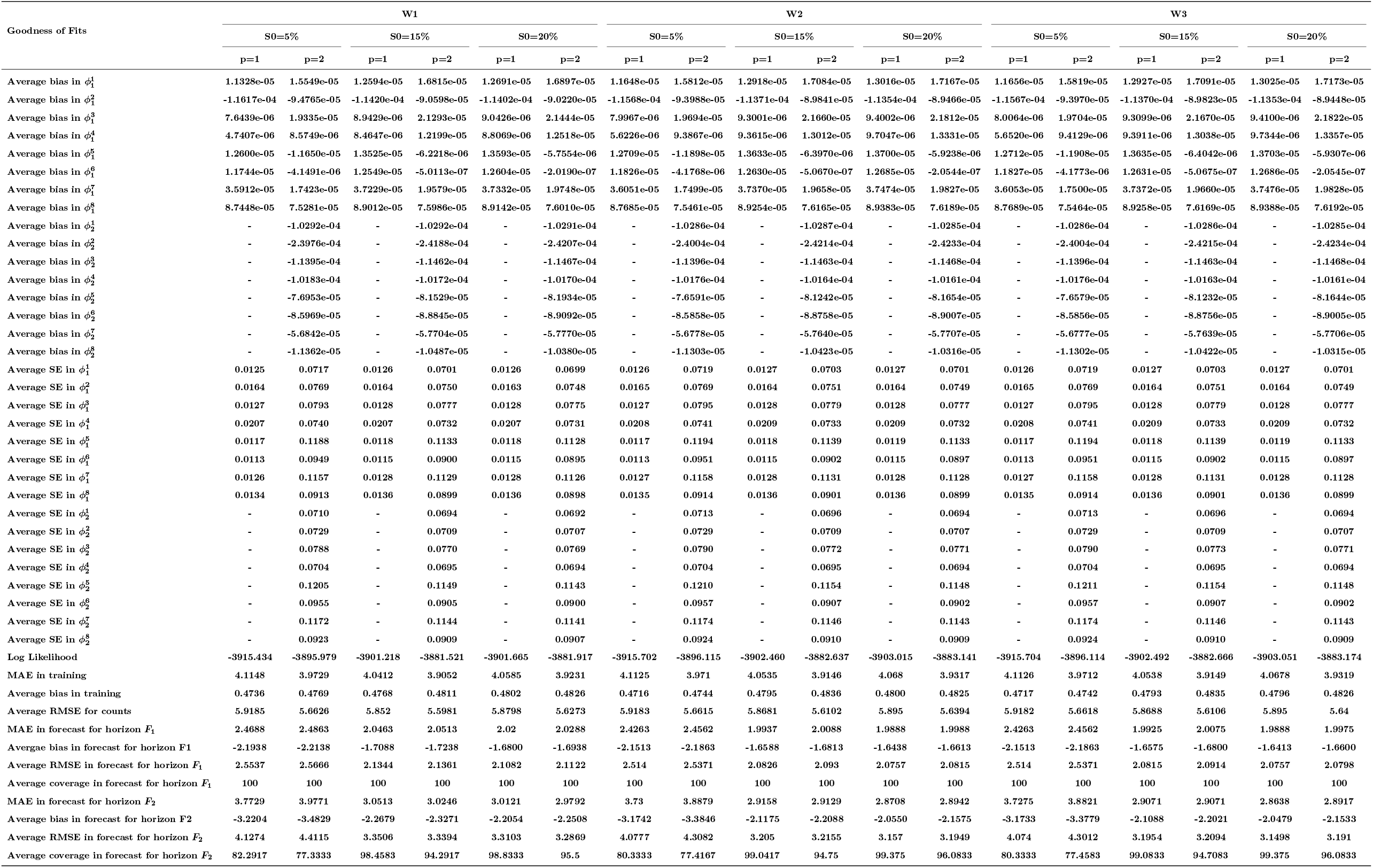
Parameter estimation and goodness of fit statistics of the proposed multi-region chain binomial model for case 3, Section 3.1.

| Goodness of Fits | W1 |  |  |  |  |  | W2 |  |  |  |  |  | W3 |  |  |  |  |  |
|---|---|---|---|---|---|---|---|---|---|---|---|---|---|---|---|---|---|---|
|  | S0=5% |  | S0=15% |  | S0=20% |  | S0=5% |  | S0=15% |  | S0=20% |  | S0=5% |  | S0=15% |  | S0=20% |  |
|  | p=1 | p=2 | p=1 | p=2 | p=1 | p=2 | p=1 | p=2 | p=1 | p=2 | p=1 | p=2 | p=1 | p=2 | p=1 | p=2 | p=1 | p=2 |
| Average bias in ϕ<sub>1</sub><sup>1</sup> | 1.1328e-05 | 1.5549e-05 | 1.2594e-05 | 1.6815e-05 | 1.2691e-05 | 1.6897e-05 | 1.1648e-05 | 1.5812e-05 | 1.2918e-05 | 1.7084e-05 | 1.3016e-05 | 1.7167e-05 | 1.1656e-05 | 1.5819e-05 | 1.2927e-05 | 1.7091e-05 | 1.3025e-05 | 1.7173e-05 |
| Average bias in ϕ<sub>1</sub><sup>2</sup> | -1.1617e-04 | -9.4765e-05 | -1.1420e-04 | -9.0598e-05 | -1.1402e-04 | -9.0220e-05 | -1.1568e-04 | -9.3988e-05 | -1.1371e-04 | -8.9841e-05 | -1.1354e-04 | -8.9466e-05 | -1.1567e-04 | -9.3970e-05 | -1.1370e-04 | -8.9823e-05 | -1.1353e-04 | -8.9448e-05 |
| Average bias in ϕ<sub>1</sub><sup>3</sup> | 7.6439e-06 | 1.9335e-05 | 8.9429e-06 | 2.1293e-05 | 9.0426e-06 | 2.1444e-05 | 7.9967e-06 | 1.9694e-05 | 9.3001e-06 | 2.1660e-05 | 9.4002e-06 | 2.1812e-05 | 8.0064e-06 | 1.9704e-05 | 9.3099e-06 | 2.1670e-05 | 9.4100e-06 | 2.1822e-05 |
| Average bias in ϕ<sub>1</sub><sup>4</sup> | 4.7407e-06 | 8.5749e-06 | 8.4647e-06 | 1.2199e-05 | 8.8069e-06 | 1.2518e-05 | 5.6226e-06 | 9.3867e-06 | 9.3615e-06 | 1.3012e-05 | 9.7047e-06 | 1.3331e-05 | 5.6520e-06 | 9.4129e-06 | 9.3911e-06 | 1.3038e-05 | 9.7344e-06 | 1.3357e-05 |
| Average bias in ϕ<sub>1</sub><sup>5</sup> | 1.2600e-05 | -1.1650e-05 | 1.3525e-05 | -6.2218e-06 | 1.3593e-05 | -5.7554e-06 | 1.2709e-05 | -1.1898e-05 | 1.3633e-05 | -6.3970e-06 | 1.3700e-05 | -5.9238e-06 | 1.2712e-05 | -1.1908e-05 | 1.3635e-05 | -6.4042e-06 | 1.3703e-05 | -5.9307e-06 |
| Average bias in ϕ<sub>1</sub><sup>6</sup> | 1.1744e-05 | -4.1491e-06 | 1.2549e-05 | -5.0113e-07 | 1.2604e-05 | -2.0190e-07 | 1.1826e-05 | -4.1768e-06 | 1.2630e-05 | -5.0670e-07 | 1.2685e-05 | -2.0544e-07 | 1.1827e-05 | -4.1773e-06 | 1.2631e-05 | -5.0675e-07 | 1.2686e-05 | -2.0545e-07 |
| Average bias in ϕ<sub>1</sub><sup>7</sup> | 3.5912e-05 | 1.7423e-05 | 3.7229e-05 | 1.9579e-05 | 3.7332e-05 | 1.9748e-05 | 3.6051e-05 | 1.7499e-05 | 3.7370e-05 | 1.9658e-05 | 3.7474e-05 | 1.9827e-05 | 3.6053e-05 | 1.7500e-05 | 3.7372e-05 | 1.9660e-05 | 3.7476e-05 | 1.9828e-05 |
| Average bias in ϕ<sub>1</sub><sup>8</sup> | 8.7448e-05 | 7.5281e-05 | 8.9012e-05 | 7.5986e-05 | 8.9142e-05 | 7.6010e-05 | 8.7685e-05 | 7.5461e-05 | 8.9254e-05 | 7.6165e-05 | 8.9383e-05 | 7.6189e-05 | 8.7689e-05 | 7.5464e-05 | 8.9258e-05 | 7.6169e-05 | 8.9388e-05 | 7.6192e-05 |
| Average bias in ϕ<sub>2</sub><sup>1</sup> | - | -1.0292e-04 | - | -1.0292e-04 | - | -1.0291e-04 | - | -1.0286e-04 | - | -1.0287e-04 | - | -1.0285e-04 | - | -1.0286e-04 | - | -1.0286e-04 | - | -1.0285e-04 |
| Average bias in ϕ<sub>2</sub><sup>2</sup> | - | -2.3976e-04 | - | -2.4188e-04 | - | -2.4207e-04 | - | -2.4004e-04 | - | -2.4214e-04 | - | -2.4233e-04 | - | -2.4004e-04 | - | -2.4215e-04 | - | -2.4234e-04 |
| Average bias in ϕ<sub>2</sub><sup>3</sup> | - | -1.1395e-04 | - | -1.1462e-04 | - | -1.1467e-04 | - | -1.1396e-04 | - | -1.1463e-04 | - | -1.1468e-04 | - | -1.1396e-04 | - | -1.1463e-04 | - | -1.1468e-04 |
| Average bias in ϕ<sub>2</sub><sup>4</sup> | - | -1.0183e-04 | - | -1.0172e-04 | - | -1.0170e-04 | - | -1.0176e-04 | - | -1.0164e-04 | - | -1.0161e-04 | - | -1.0176e-04 | - | -1.0163e-04 | - | -1.0161e-04 |
| Average bias in ϕ<sub>2</sub><sup>5</sup> | - | -7.6953e-05 | - | -8.1529e-05 | - | -8.1934e-05 | - | -7.6591e-05 | - | -8.1242e-05 | - | -8.1654e-05 | - | -7.6579e-05 | - | -8.1232e-05 | - | -8.1644e-05 |
| Average bias in ϕ<sub>2</sub><sup>6</sup> | - | -8.5969e-05 | - | -8.8845e-05 | - | -8.9092e-05 | - | -8.5858e-05 | - | -8.8758e-05 | - | -8.9007e-05 | - | -8.5856e-05 | - | -8.8756e-05 | - | -8.9005e-05 |
| Average bias in ϕ<sub>2</sub><sup>7</sup> | - | -5.6842e-05 | - | -5.7704e-05 | - | -5.7770e-05 | - | -5.6778e-05 | - | -5.7640e-05 | - | -5.7707e-05 | - | -5.6777e-05 | - | -5.7639e-05 | - | -5.7706e-05 |
| Average bias in ϕ<sub>2</sub><sup>8</sup> | - | -1.1362e-05 | - | -1.0487e-05 | - | -1.0380e-05 | - | -1.1303e-05 | - | -1.0423e-05 | - | -1.0316e-05 | - | -1.1302e-05 | - | -1.0422e-05 | - | -1.0315e-05 |
| Average SE in ϕ<sub>1</sub><sup>1</sup> | 0.0125 | 0.0717 | 0.0126 | 0.0701 | 0.0126 | 0.0699 | 0.0126 | 0.0719 | 0.0127 | 0.0703 | 0.0127 | 0.0701 | 0.0126 | 0.0719 | 0.0127 | 0.0703 | 0.0127 | 0.0701 |
| Average SE in ϕ<sub>1</sub><sup>2</sup> | 0.0164 | 0.0769 | 0.0164 | 0.0750 | 0.0163 | 0.0748 | 0.0165 | 0.0769 | 0.0164 | 0.0751 | 0.0164 | 0.0749 | 0.0165 | 0.0769 | 0.0164 | 0.0751 | 0.0164 | 0.0749 |
| Average SE in ϕ<sub>1</sub><sup>3</sup> | 0.0127 | 0.0793 | 0.0128 | 0.0777 | 0.0128 | 0.0775 | 0.0127 | 0.0795 | 0.0128 | 0.0779 | 0.0128 | 0.0777 | 0.0127 | 0.0795 | 0.0128 | 0.0779 | 0.0128 | 0.0777 |
| Average SE in ϕ<sub>1</sub><sup>4</sup> | 0.0207 | 0.0740 | 0.0207 | 0.0732 | 0.0207 | 0.0731 | 0.0208 | 0.0741 | 0.0209 | 0.0733 | 0.0209 | 0.0732 | 0.0208 | 0.0741 | 0.0209 | 0.0733 | 0.0209 | 0.0732 |
| Average SE in ϕ<sub>1</sub><sup>5</sup> | 0.0117 | 0.1188 | 0.0118 | 0.1133 | 0.0118 | 0.1128 | 0.0117 | 0.1194 | 0.0118 | 0.1139 | 0.0119 | 0.1133 | 0.0117 | 0.1194 | 0.0118 | 0.1139 | 0.0119 | 0.1133 |
| Average SE in ϕ<sub>1</sub><sup>6</sup> | 0.0113 | 0.0949 | 0.0115 | 0.0900 | 0.0115 | 0.0895 | 0.0113 | 0.0951 | 0.0115 | 0.0902 | 0.0115 | 0.0897 | 0.0113 | 0.0951 | 0.0115 | 0.0902 | 0.0115 | 0.0897 |
| Average SE in ϕ<sub>1</sub><sup>7</sup> | 0.0126 | 0.1157 | 0.0128 | 0.1129 | 0.0128 | 0.1126 | 0.0127 | 0.1158 | 0.0128 | 0.1131 | 0.0128 | 0.1128 | 0.0127 | 0.1158 | 0.0128 | 0.1131 | 0.0128 | 0.1128 |
| Average SE in ϕ<sub>1</sub><sup>8</sup> | 0.0134 | 0.0913 | 0.0136 | 0.0899 | 0.0136 | 0.0898 | 0.0135 | 0.0914 | 0.0136 | 0.0901 | 0.0136 | 0.0899 | 0.0135 | 0.0914 | 0.0136 | 0.0901 | 0.0136 | 0.0899 |
| Average SE in ϕ<sub>2</sub><sup>1</sup> | - | 0.0710 | - | 0.0694 | - | 0.0692 | - | 0.0713 | - | 0.0696 | - | 0.0694 | - | 0.0713 | - | 0.0696 | - | 0.0694 |
| Average SE in ϕ<sub>2</sub><sup>2</sup> | - | 0.0729 | - | 0.0709 | - | 0.0707 | - | 0.0729 | - | 0.0709 | - | 0.0707 | - | 0.0729 | - | 0.0709 | - | 0.0707 |
| Average SE in ϕ<sub>2</sub><sup>3</sup> | - | 0.0788 | - | 0.0770 | - | 0.0769 | - | 0.0790 | - | 0.0772 | - | 0.0771 | - | 0.0790 | - | 0.0773 | - | 0.0771 |
| Average SE in ϕ<sub>2</sub><sup>4</sup> | - | 0.0704 | - | 0.0695 | - | 0.0694 | - | 0.0704 | - | 0.0695 | - | 0.0694 | - | 0.0704 | - | 0.0695 | - | 0.0694 |
| Average SE in ϕ<sub>2</sub><sup>5</sup> | - | 0.1205 | - | 0.1149 | - | 0.1143 | - | 0.1210 | - | 0.1154 | - | 0.1148 | - | 0.1211 | - | 0.1154 | - | 0.1148 |
| Average SE in ϕ<sub>2</sub><sup>6</sup> | - | 0.0955 | - | 0.0905 | - | 0.0900 | - | 0.0957 | - | 0.0907 | - | 0.0902 | - | 0.0957 | - | 0.0907 | - | 0.0902 |
| Average SE in ϕ<sub>2</sub><sup>7</sup> | - | 0.1172 | - | 0.1144 | - | 0.1141 | - | 0.1174 | - | 0.1146 | - | 0.1143 | - | 0.1174 | - | 0.1146 | - | 0.1143 |
| Average SE in ϕ<sub>2</sub><sup>8</sup> | - | 0.0923 | - | 0.0909 | - | 0.0907 | - | 0.0924 | - | 0.0910 | - | 0.0909 | - | 0.0924 | - | 0.0910 | - | 0.0909 |
| Log Likelihood | -3915.434 | -3895.979 | -3901.218 | -3881.521 | -3901.665 | -3881.917 | -3915.702 | -3896.115 | -3902.460 | -3882.637 | -3903.015 | -3883.141 | -3915.704 | -3896.114 | -3902.492 | -3882.666 | -3903.051 | -3883.174 |
| MAE in training | 4.1148 | 3.9729 | 4.0412 | 3.9052 | 4.0585 | 3.9231 | 4.1125 | 3.971 | 4.0535 | 3.9146 | 4.068 | 3.9317 | 4.1126 | 3.9712 | 4.0538 | 3.9149 | 4.0678 | 3.9319 |
| Average bias in training | 0.4736 | 0.4769 | 0.4768 | 0.4811 | 0.4802 | 0.4826 | 0.4716 | 0.4744 | 0.4795 | 0.4836 | 0.4800 | 0.4825 | 0.4717 | 0.4742 | 0.4793 | 0.4835 | 0.4796 | 0.4826 |
| Average RMSE for counts | 5.9185 | 5.6626 | 5.852 | 5.5981 | 5.8798 | 5.6273 | 5.9183 | 5.6615 | 5.8681 | 5.6102 | 5.895 | 5.6394 | 5.9182 | 5.6618 | 5.8688 | 5.6106 | 5.895 | 5.64 |
| MAE in forecast for horizon F<sub>1</sub> | 2.4688 | 2.4863 | 2.0463 | 2.0513 | 2.02 | 2.0288 | 2.4263 | 2.4562 | 1.9937 | 2.0088 | 1.9888 | 1.9988 | 2.4263 | 2.4562 | 1.9925 | 2.0075 | 1.9888 | 1.9975 |
| Avergae bias in forecast for horizon F1 | -2.1938 | -2.2138 | -1.7088 | -1.7238 | -1.6800 | -1.6938 | -2.1513 | -2.1863 | -1.6588 | -1.6813 | -1.6438 | -1.6613 | -2.1513 | -2.1863 | -1.6575 | -1.6800 | -1.6413 | -1.6600 |
| Average RMSE in forecast for horizon F<sub>1</sub> | 2.5537 | 2.5666 | 2.1344 | 2.1361 | 2.1082 | 2.1122 | 2.514 | 2.5371 | 2.0826 | 2.093 | 2.0757 | 2.0815 | 2.514 | 2.5371 | 2.0815 | 2.0914 | 2.0757 | 2.0798 |
| Average coverage in forecast for horizon F<sub>1</sub> | 100 | 100 | 100 | 100 | 100 | 100 | 100 | 100 | 100 | 100 | 100 | 100 | 100 | 100 | 100 | 100 | 100 | 100 |
| MAE in forecast for horizon F<sub>2</sub> | 3.7729 | 3.9771 | 3.0513 | 3.0246 | 3.0121 | 2.9792 | 3.73 | 3.8879 | 2.9158 | 2.9129 | 2.8708 | 2.8942 | 3.7275 | 3.8821 | 2.9071 | 2.9071 | 2.8638 | 2.8917 |
| Average bias in forecast for horizon F2 | -3.2204 | -3.4829 | -2.2679 | -2.3271 | -2.2054 | -2.2508 | -3.1742 | -3.3846 | -2.1175 | -2.2088 | -2.0550 | -2.1575 | -3.1733 | -3.3779 | -2.1088 | -2.2021 | -2.0479 | -2.1533 |
| Average RMSE in forecast for horizon F<sub>2</sub> | 4.1274 | 4.4115 | 3.3506 | 3.3394 | 3.3103 | 3.2869 | 4.0777 | 4.3082 | 3.205 | 3.2155 | 3.157 | 3.1949 | 4.074 | 4.3012 | 3.1954 | 3.2094 | 3.1498 | 3.191 |
| Average coverage in forecast for horizon F<sub>2</sub> | 82.2917 | 77.3333 | 98.4583 | 94.2917 | 98.8333 | 95.5 | 80.3333 | 77.4167 | 99.0417 | 94.75 | 99.375 | 96.0833 | 80.3333 | 77.4583 | 99.0833 | 94.7083 | 99.375 | 96.0833 |

We also compared the performance of our proposed approach with the HHH model for cases 1 and 2. HHH model was implemented using the hhh4 function of the surveillance package in R (Meyer et al. [2017]). Tables 4 and Table 5 gives the comparison results for cases 1 and 2, respectively. From these two tables we note that the proposed multi-region chain binomial model outperforms the HHH model for both prediction and forecasting of counts when the susceptiple pool is smaller. The underlying Poisson assumption for the HHH model seems to be not working as well when the susceptible pool is smaller. Thus, we may infer that the proposed framework is expected to perfrom better in settings where susceptible depletion is substantial relative to population size, such as smaller populations or localized outbreaks.

**Table 4:** Comparison between the proposed multi-region chain binomial model (MCB) and HHH model for case 1, Section 3.1. The GOF metrics are averaged over the regions.

| **GOF Metric** | **MCB** | **HHH** |
|---|---|---|
| Average bias in training | 4.4188 | 5.3843 |
| Average RMSE in training | 6.3107 | 8.0397 |
| Average bias in forecast for horizon F1 | 2.4046 | 1.2786 |
| Average RMSE in forecast for horizon F1 | 2.5583 | 1.4340 |
| Average coverage in forecast for horizon F1 | 100.0000 | 100.0000 |
| Average bias in forecast for horizon F2 | 3.3646 | 1.9333 |
| Average RMSE in forecast for horizon F2 | 3.8099 | 2.2439 |
| Average coverage in forecast for horizon F2 | 97.8667 | 100.0000 |

**Table 5:** Comparison between the proposed multi-region chain binomial model (MCB) and HHH model for case 2, Section 3.1. The GOF metrics are averaged over the regions.

| **GOF Metric** | **MCB** | **HHH** |
|---|---|---|
| Average bias in training | 1.1781 | 1.3420 |
| Average RMSE in training | 1.5392 | 1.7735 |
| Average bias in forecast for horizon F1 | 2.1430 | 4.8240 |
| Average RMSE in forecast for horizon F1 | 2.3962 | 5.1316 |
| Average coverage in forecast for horizon F1 | 96.2000 | 92.0000 |
| Average bias in forecast for horizon F2 | 2.0220 | 5.2030 |
| Average RMSE in forecast for horizon F2 | 2.4579 | 5.6613 |
| Average coverage in forecast for horizon F2 | 89.7000 | 89.6667 |

**Table 6:** True parameter values for Cases 1, 2 and 3, Section 3.1.

|  | **Case 1** |  |  |  |  | **Case 2** |  |  |  |  | **Case 3** |  |  |  |  |  |  |  |
|---|---|---|---|---|---|---|---|---|---|---|---|---|---|---|---|---|---|---|
| **Parameter** | **R1** | **R2** | **R3** | **R4** | **R5** | **R1** | **R2** | **R3** | **R4** | **R5** | **R1** | **R2** | **R3** | **R4** | **R5** | **R6** | **R7** | **R8** |
| **β<sub>0</sub>** | 0.7000 | 0.7000 | 0.7000 | 0.7000 | 0.7000 | 0.7000 | 0.7000 | 0.7000 | 0.7000 | 0.7000 | 0.7000 | 0.7000 | 0.7000 | 0.7000 | 0.7000 | 0.7000 | 0.7000 | 0.7000 |
| **ϕ<sub>1</sub>** | 0.0001 | 0.0002 | 0.0001 | 0.0001 | 0.0001 | 0.0111 | 0.0121 | 0.0115 | 0.0110 | 0.0114 | 0.0001 | 0.0002 | 0.0001 | 0.0001 | 0.0001 | 0.0001 | 0.0002 | 0.0001 |
| **ϕ<sub>2</sub>** | 0.0001 | 0.0002 | 0.0001 | 0.0001 | 0.0001 | 0.0086 | 0.0109 | 0.0090 | 0.0085 | 0.0064 | 0.0001 | 0.0002 | 0.0001 | 0.0001 | 0.0001 | 0.0001 | 0.0002 | 0.0001 |
| **β<sub>1</sub>** | 0.0389 | 0.0233 | 0.0373 | 0.0279 | 0.0515 | 0.0389 | 0.0233 | 0.0373 | 0.0279 | 0.0515 | 0.0389 | 0.0233 | 0.0373 | 0.0279 | 0.0515 | 0.0515 | 0.0415 | 0.0415 |
| **β<sub>2</sub>** | 0.1527 | 0.1907 | 0.1943 | 0.1836 | 0.0344 | 0.1527 | 0.1907 | 0.1943 | 0.1836 | 0.0344 | 0.1527 | 0.1907 | 0.1943 | 0.1836 | 0.0344 | 0.0344 | 0.0144 | 0.0144 |

### 3.2 Simulation Study 2

Instead of using equations 1 and 3 to generate the data, we used the MSIR model for data generation. This was done to check if we can apply our proposed approach to a fundamentally different data generation process and still get good prediction and forecasting results. We assume here that interventions like vaccination are available for controlling the disease. Suppose the disease being modeled is the biweekly counts of measles, which is a highly contagious, vaccine-preventable viral disease. The World Health Organization recommends administering two doses of measles vaccines at 9-12 months and 16-24 months to control and eliminate the disease. Measles have been modeled previously by various researchers, namely, Grenfell et al. [2001], Chen et al. [2012], Thakkar et al. [2019].

The biweekly counts were generated using the following susceptible-infection equations of the MSIR model. It is assumed that there are 3 spatially connected regions and 155 biweekly incidences are generated using the following equations,

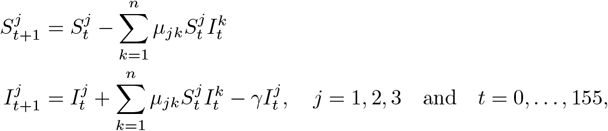

where the recovery rate, *γ*, is assumed to be 1/14 day^−1^ and *µ*_*jk*_’s are the disease transmission rates between two locations *j* and *k*. The transmission rates are taken to be influenced by region-specific factors such as the number of susceptibles and annual seasonality,

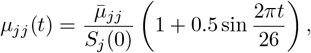

here 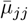 are the region-specific mean transmission rates. We model the *µ*_*jk*_’s to depend on the distance between two spatial regions *j* and *k* as: *µ*_*jk*_(*t*) = *µ*_*jj*_(*t*)*w*_*jk*_, where *w*_*jk*_’s are spatial weights.

A spatial weight matrix, *W* = ((*w*_*jk*_)),

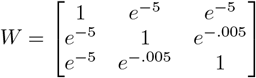

is used, which shows that regions 2 and 3 are closer to each other and equally distanced from region 1. We work with the following parameters:

- 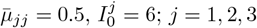 and *N*^*j*^ = 10^6^; 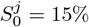 of *N*^*j*^, *∀j*.

As interventions such as vaccination strategies play an important role in measles control, we also introduce a vaccinated birth cohort as discussed in Section 2.3. The susceptible population in the equations of the MSIR model is adjusted as: 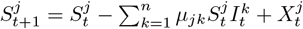 where

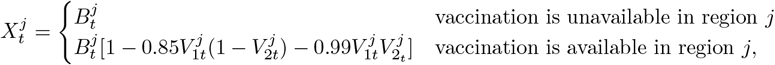

The birth cohort was chosen to remain constant over time 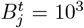, *∀j*. Thus, for a region *j* where vaccination is available, 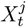 denotes the birth cohort who remains unvaccinated, thus adding to the susceptible population of the region.

We consider a scenario where we assume that vaccination is available in Region 3 only. The coverages of the two doses are, 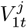 increasing linearly from 0.70 to 0.90 and 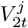 increasing linearly from 0.40 to 0.70 over the biweeks (t).

We performed 100 simulations and the proposed multi-region chain binomial model is fitted to the data. In Table 7 we present the results of fitting our model to the data generated from the MSIR model with vaccination in region 3. Note that the average coverage for one month forecasting horizon is over 93%, which drops to 75% for the three months horizon. We can thus infer that our proposed approach performs reasonably well when the disease data is generated from the MSIR model.

**Table 7:** Goodness of fit statistics for the simulation corresponding to section 3.2.

| Goodness of Fits | W1 |  |  |  |  |  | W2 |  |  |  |  |  | W3 |  |  |  |  |  |
|---|---|---|---|---|---|---|---|---|---|---|---|---|---|---|---|---|---|---|
|  | S0=5% |  | S0=15% |  | S0=20% |  | S0=5% |  | S0=15% |  | S0=20% |  | S0=5% |  | S0=15% |  | S0=20% |  |
|  | p=1 | p=2 | p=1 | p=2 | p=1 | p=2 | p=1 | p=2 | p=1 | p=2 | p=1 | p=2 | p=1 | p=2 | p=1 | p=2 | p=1 | p=2 |
| Average MAE for counts | 4.7371 | 3.3842 | 5.1254 | 3.6159 | 5.1664 | 3.6437 | 4.9824 | 3.5868 | 5.3662 | 3.8208 | 5.4074 | 3.8492 | 4.5364 | 3.2146 | 4.9266 | 3.4487 | 4.9681 | 3.4781 |
| Average RMSE for counts | 10.0571 | 6.3837 | 11.0801 | 7.0220 | 11.1895 | 7.0998 | 10.5334 | 6.7635 | 11.5443 | 7.3942 | 11.6520 | 7.4703 | 9.6771 | 6.0704 | 10.7059 | 6.7144 | 10.8143 | 6.7944 |
| Average MAE in forecast for horizon F1 | 2.3950 | 2.3950 | 2.4783 | 2.4783 | 2.4800 | 2.4800 | 2.4467 | 2.4467 | 2.5333 | 2.5333 | 2.5417 | 2.5417 | 2.3617 | 2.3617 | 2.4250 | 2.4250 | 2.4317 | 2.4317 |
| Average RMSE in forecast for horizon F1 | 2.4388 | 2.4388 | 2.5245 | 2.5245 | 2.5251 | 2.5251 | 2.4864 | 2.4864 | 2.5770 | 2.5770 | 2.5862 | 2.5862 | 2.4078 | 2.4078 | 2.4719 | 2.4719 | 2.4818 | 2.4818 |
| Average coverage in forecast for horizon F1 | 95.6667 | 95.6667 | 93.1667 | 93.1667 | 93.1667 | 93.1667 | 95.6667 | 95.6667 | 92.6667 | 92.6667 | 92.6667 | 92.6667 | 95.8333 | 95.8333 | 93.8333 | 93.8333 | 93.3333 | 93.3333 |
| Average MAE in forecast for horizon F2 | 12.7944 | 12.7944 | 13.5844 | 13.5844 | 13.6867 | 13.6867 | 13.2400 | 13.2400 | 14.1317 | 14.1317 | 14.2228 | 14.2228 | 12.4044 | 12.4044 | 13.1378 | 13.1378 | 13.2350 | 13.2350 |
| Average RMSE in forecast for horizon F2 | 14.7650 | 14.7650 | 15.6848 | 15.6848 | 15.8093 | 15.8093 | 15.2982 | 15.2982 | 16.3595 | 16.3595 | 16.4690 | 16.4690 | 14.2696 | 14.2696 | 15.1345 | 15.1345 | 15.2455 | 15.2455 |
| Average coverage in forecast for horizon F2 | 77.3889 | 77.3889 | 75.9444 | 76.0556 | 76.2222 | 76.0556 | 77.0000 | 76.8333 | 75.3333 | 75.3333 | 75.8889 | 75.8333 | 78.6111 | 78.6667 | 76.7778 | 77.3333 | 76.9444 | 76.6667 |

## 4 Real Data Example: Analysis of Measles Counts in UK

To illustrate our proposed model on real data, we fitted it to biweekly case reports of measles recorded between the years 1944-1966 from seven urban cities, Birmingham, Coventry, Leicester, Derby, Nottingham, Wolverhampton and Walsall, in the United Kingdom (UK). The UK measles data set has previously been analyzed by various authors, namely (Grenfell et al. [2001], Xia et al. [2004], Madden et al. [2024]). We chose the seven cities due to their spatial proximities (distance) on the UK map. From an initial spatial correlation analysis, a Moran’s-I value (Anselin [1995]) of 3.0277 and a p-value of 0.0012 (significant at the 5% level) were observed, implying existence of spatial correlation among the disease counts (averaged over time) of the seven cities. During the period 1944 to 1966, these seven English Midlands cities (see Figure 1) were connected through a complex and evolving public transport network Eye [2016] that facilitated the frequent movement of individuals among the cities. The railway system served as the primary inter-city connector, with Birmingham operating as the central hub thus, justifying the highest case counts of Birmingham. For the location of the cities and the strength of the public transport connection between them, see Figure 1, while for distances between cities, see Table 8. From Figure 1, as Birmingham is the most strongly connected to all cities, outbreaks in all cities seem to be synchronized or follow. These seven cities also differ in their population status, with Birmingham having the highest population. We divide the cities that exclude Birmingham into two groups (high/low) according to their population densities and plot the measles counts to compare the incidences and pattern with Birmingham in Figure 2.

**Table 8:**
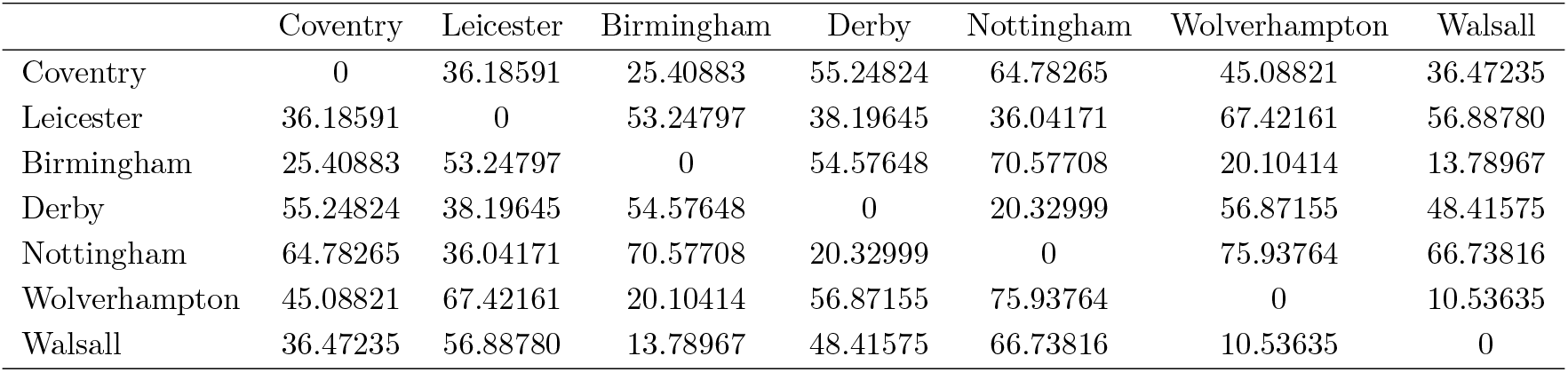
Distance in km among the seven UK cities in Section 4.

**Figure 1:**
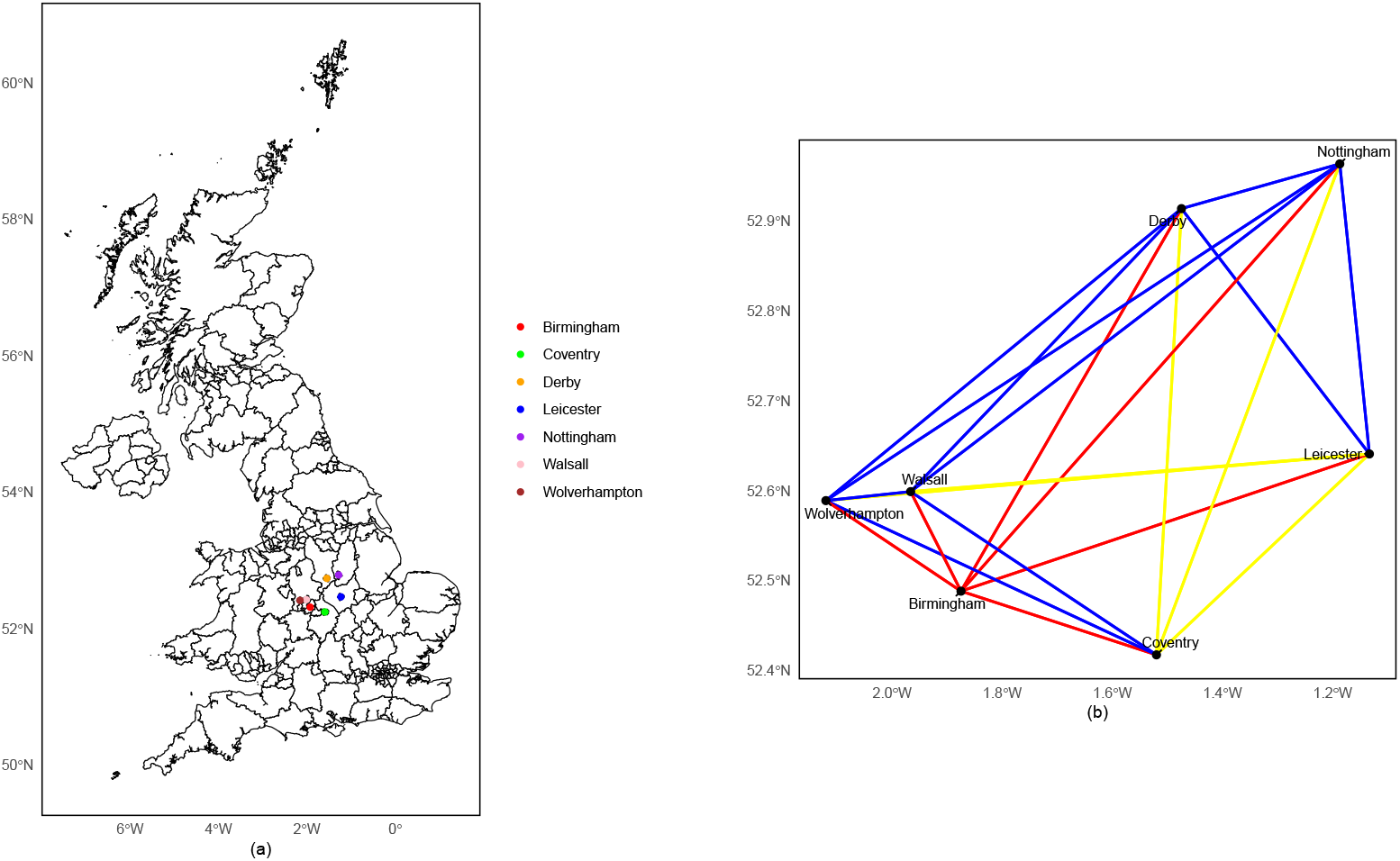
Left panel (a) of the figure shows the location of the seven cities in the UK map and the right panel (b) shows the strength of connection between the seven UK cities. In (b), the red lines represent strong connections, blue lines represent moderate connections and yellow lines represent weak connections, corresponding to Section 4.

**Figure 2:**
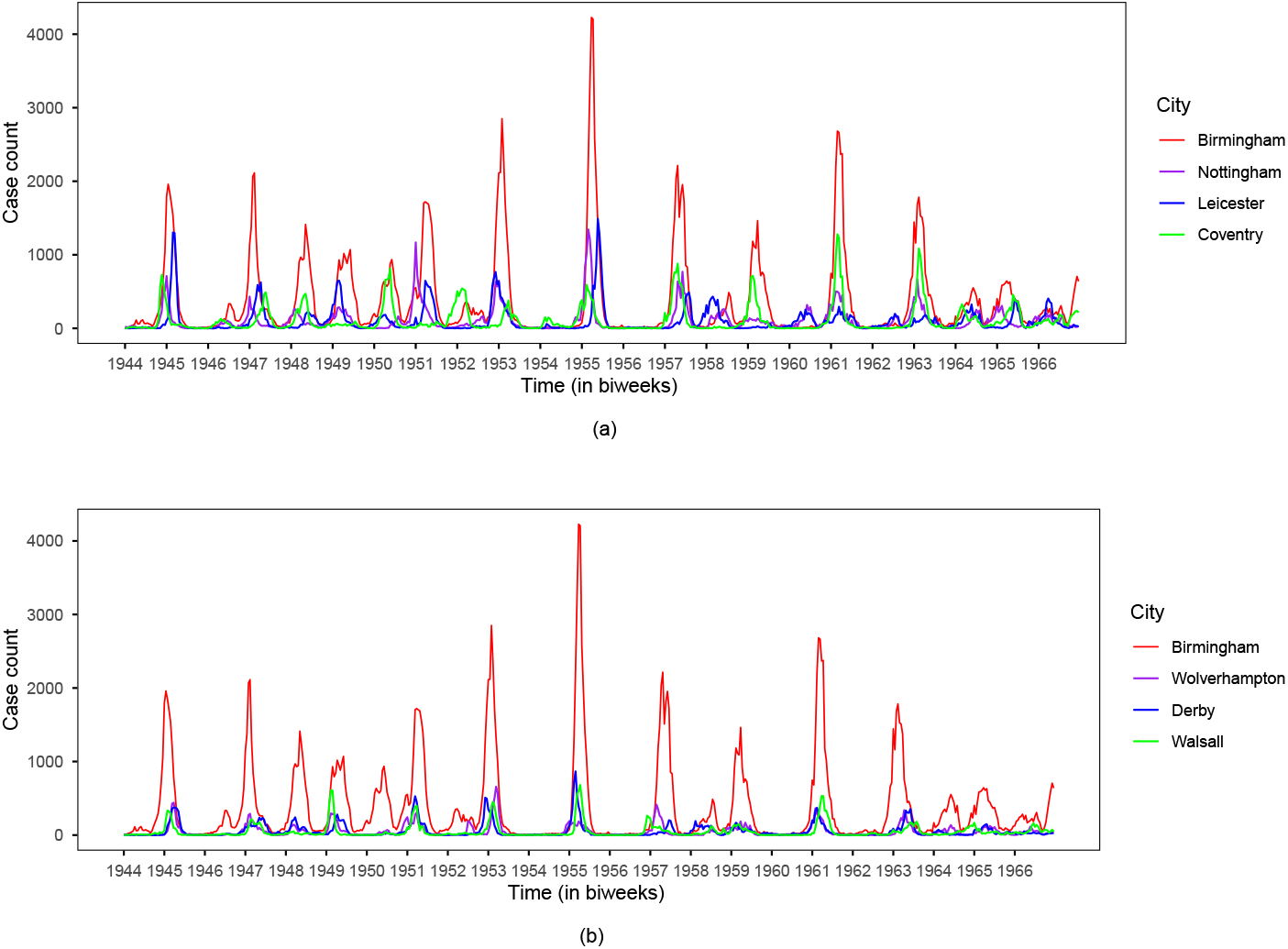
Outbreak comparisons between Birmingham and other cities on the basis of raw incidences in Section 4. The cities in (a) are the high population density cities while in (b) we have the low population density cities, corresponding to Section 4.

The measles data was divided into a training set and a test set. The training sets were chosen as the period 1945 (January) to 1966 (June) and thereafter were the test data. Using various forms of equation 3, our objective was to select the best model to understand the spatial coupling between these seven UK cities, study the transmission within and between cities through 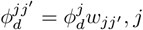, *j*^*′*^ = 1, …, 7, and the region specific trend, seasonality and covariates which affect the spread of the disease. For this we studied long term trend effect, seasonality (annual and semi-annual) and covariates, biweekly birth counts and the 1946 baby boom effect in the model. To select the lag order *d*, we tested different values of p ranging from 1 to 7, and ranked the models according to their BIC values, the lowest BIC (1286.25) was obtained for p=1. We checked various forms of *g*(*·*) in equation 3; and, the best fitting results were obtained for the logarithm function. To find 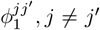, we studied the relation 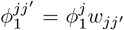 for two different weight functions, 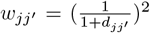, and 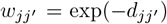. The best models chosen based on the various GOFs and the training set for each of the 7 cities are described in Table 9.

**Table 9:**
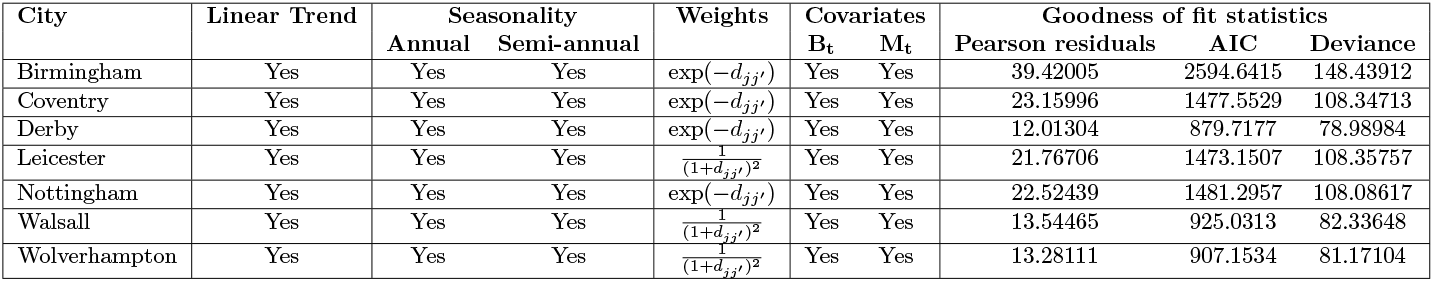
Description of best model for each of the seven UK cities in Section 4. The Goodness of fit statistics, reported in the last column, are best among competing models.

Incidence counts across all spatial units constitute a multivariate time series with inter-dependencies; however, conditional on 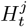, the incidence time series for each region can be treated as independent as shown in equation 1. Consequently, we applied the GLM function from the base stats package in R to each city separately, using the model specification described above. Estimates of model parameters, standard errors, and p-values for each of the seven cities are listed in Table 10. From Table 10, we observe an increasing trend in the incidence of measles in all cities. Strong seasonal patterns, both semi-annual and annual, are evident across all locations, indicating periodic oscillations in measles transmission. The covariates related to demographic dynamics, specifically live births and the post-World War-II baby boom, demonstrate significant associations with the incidence of measles. The region-specific parameter, 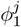, modeling the effect of disease transmission within the city was significant in all cities. We see that the odds of infection increase by a factor of 2.50 for Birmingham, 2.49 for Nottingham, 2.39 for Leicester, 2.43 for Coventry, 2.29 for Wolverhampton, 2.41 for Derby, 2.48 for Walsall.

**Table 10:** Parameter estimates, standard errors and p-values of the models fitted to the measles counts from all the 7 UK cities in Section 4.

| **Variable** | **Estimate** | **SE** | **p-value** |
|---|---|---|---|
| (a) Birmingham |  |  |  |
| Intercept | −13.2100 | 0.0314 | <.001 |
| ϕ<sub>1</sub><sup>B</sup> | 0.9195 | 0.0026 | <.001 |
| Bi-weekly Birth Counts | 0.0003 | 0.0000 | <.001 |
| Annual Seasonality | 0.1238 | 0.0039 | <.001 |
| Semi-annual Seasonality | 0.1943 | 0.0046 | <.001 |
| Trend | 0.0004 | 0.0000 | <.001 |
| Baby Boom Effect | 0.0839 | 0.0069 | <.001 |
| (b) Coventry |  |  |  |
| Intercept | −12.1300 | 0.0349 | <.001 |
| ϕ<sub>1</sub><sup>C</sup> | 0.8880 | 0.0041 | <.001 |
| Bi-weekly Birth Counts | −0.0012 | 0.0002 | <.001 |
| Annual Seasonality | 0.0574 | 0.0065 | <.001 |
| Semi-annual Seasonality | 0.1527 | 0.0084 | <.001 |
| Trend | 0.0004 | 0.0000 | <.001 |
| Baby Boom Effect | 0.0631 | 0.0137 | <.001 |
| (c) Derby |  |  |  |
| Intercept | −10.9500 | 0.0731 | <.001 |
| ϕ<sub>1</sub><sup>D</sup> | 0.8819 | 0.0061 | <.001 |
| Bi-weekly Birth Counts | 0.0025 | 0.0006 | <.001 |
| Annual Seasonality | 0.0690 | 0.0099 | <.001 |
| Semi-annual Seasonality | 0.1836 | 0.0113 | <.001 |
| Trend | 0.0003 | 0.0000 | <.001 |
| Baby Boom Effect | 0.0731 | 0.0187 | <.001 |
| (d) Leicester |  |  |  |
| Intercept | −11.3000 | 0.0599 | <.001 |
| ϕ<sub>1</sub><sup>L</sup> | 0.8729 | 0.0040 | <.001 |
| Bi-weekly Birth Counts | 0.0029 | 0.0003 | <.001 |
| Annual Seasonality | 0.1925 | 0.0627 | <.001 |
| Semi-annual Seasonality | 0.1907 | 0.0073 | <.001 |
| Trend | 0.0002 | 0.0000 | <.001 |
| Baby Boom Effect | 0.0600 | 0.0132 | <.001 |
| (e) Nottingham |  |  |  |
| Intercept | −11.8800 | 0.0599 | <.001 |
| ϕ<sub>1</sub><sup>N</sup> | 0.9146 | 0.0044 | <.001 |
| Bi-weekly Birth Counts | 0.0015 | 0.0002 | <.001 |
| Annual Seasonality | 0.0966 | 0.0066 | <.001 |
| Semi-annual Seasonality | 0.0344 | 0.0075 | <.001 |
| Trend | 0.0005 | 0.0000 | <.001 |
| Baby Boom Effect | 0.0974 | 0.0131 | <.001 |
| (f) Walsall |  |  |  |
| Intercept | −11.0800 | 0.0682 | <.001 |
| ϕ<sub>1</sub><sup>W</sup> | 0.9100 | 0.0062 | <.001 |
| Bi-weekly Birth Counts | 0.0016 | 0.0007 | 0.0223 |
| Annual Seasonality | 0.1912 | 0.0126 | <.001 |
| Semi-annual Seasonality | 0.1807 | 0.0128 | <.001 |
| Trend | 0.0007 | 0.0000 | <.001 |
| Baby Boom Effect | 0.0240 | 0.0205 | 0.2428 |
| (g) Wolverhampton |  |  |  |
| Intercept | −10.8500 | 0.0535 | <.001 |
| ϕ<sub>1</sub><sup>WO</sup> | 0.8304 | 0.0065 | <.001 |
| Bi-weekly Birth Counts | 0.0028 | 0.0004 | <.001 |
| Annual Seasonality | 0.1446 | 0.0109 | <.001 |
| Semi-annual Seasonality | 0.3037 | 0.0129 | <.001 |
| Trend | 0.0004 | 0.0000 | <.001 |
| Baby Boom Effect | 0.0108 | 0.0185 | 0.5580 |

Short- and long term forecasting was performed in the range of 1 month, 6 months, and 12 months for the test data along with 95% prediction intervals using the method described in section 2.4. The accuracy of the forecast is evaluated using the RMSE and MAE in three forecasting horizons. We note that both in-sample and forecast performance was the worst for Birmingham, the city with the highest population size (see Figure 3). HDR prediction intervals for both RMSE and MAE obtained using the methodology in Section 2.4 are also plotted in Figure 3.

**Figure 3:**
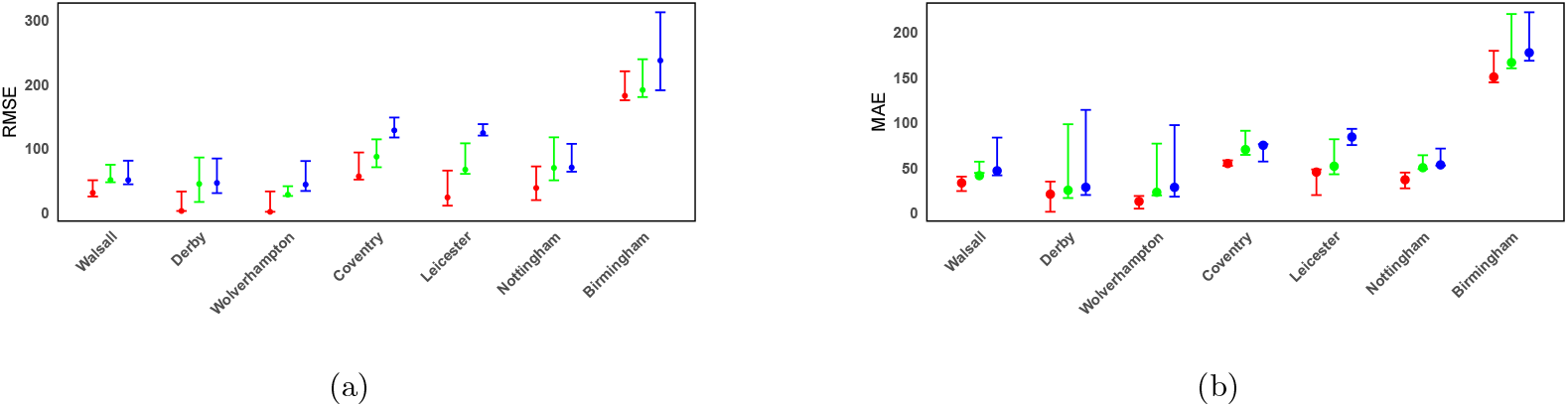
Forecasting performance for the seven cities. The left panel (a) shows the RMSE and right panel (b) shows the MAE of 1 month (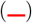), 6 months (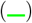) and, 12 months (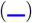), respectively. Each dot indicates the RMSE or MAE value, and the surrounding whiskers represent the confidence interval of the RMSE or MAE. Cities are ordered on the x-axis according to their population, Section 4.

## 5 Analysis of Measles Counts in West Bengal

Since measles is a vaccine-preventable disease, its outbreaks can be largely controlled through immunization. In most parts of the world, measles is now close to eradication due to widespread vaccination and cumulative immunity developed through both vaccination and prior infection. West Bengal, a state in India with an estimated population of 99.36 million, introduced the first dose of the measles-containing vaccine (MCV1) in 1985 and the second dose (MCV2) in 2010 as part of India’s measles elimination program.

We extracted monthly aggregated measles cases, and immunization information for first and second doses of measles in all districts of West Bengal from March 2014 to April 2020 from the Health Management Information System (HMIS) database, which is publicly available and downloadable at https://hmis.mohfw.gov.in/#!/standardReports. Although the state currently has 23 districts, it underwent several administrative boundary changes during the study period. The Barddhaman district was divided into Purba Barddhaman and Paschim Barddhaman, and two new districts, Jhargram and Kalimpong, were created. In addition, Alipurduar was separated from Jalpaiguri and established as an independent district in 2015. For district specific measles cases and vaccination coverages, see Figures 4 and 5.

**Figure 4:**
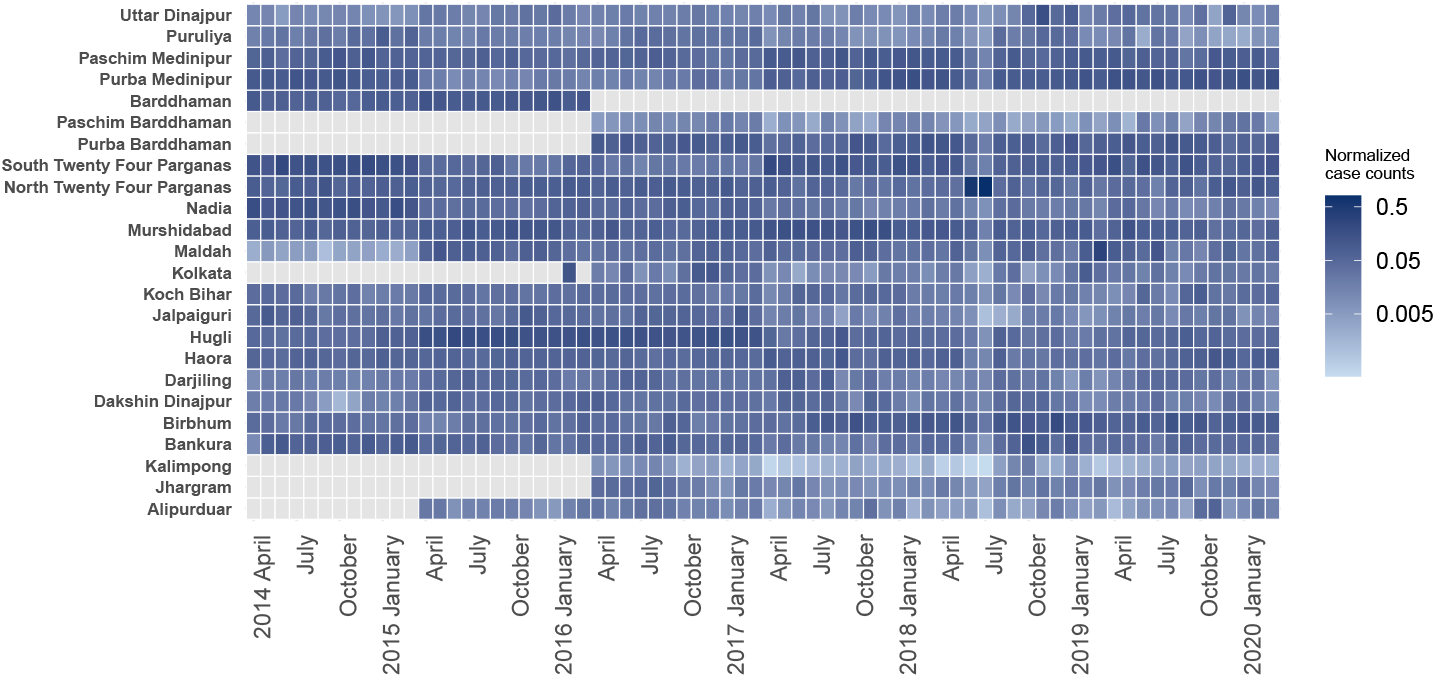
Normalized measles incidence rates (in monthly scale) for districts in West Bengal (2014-2020) corresponding to Section 5. The grey color indicates either districts that did not exist at the time (e.g., Jhargram, Kalimpong, Alipurduar, Purba Barddhaman and Paschim Barddhaman) or areas with missing data (e.g., Kolkata).

**Figure 5:**
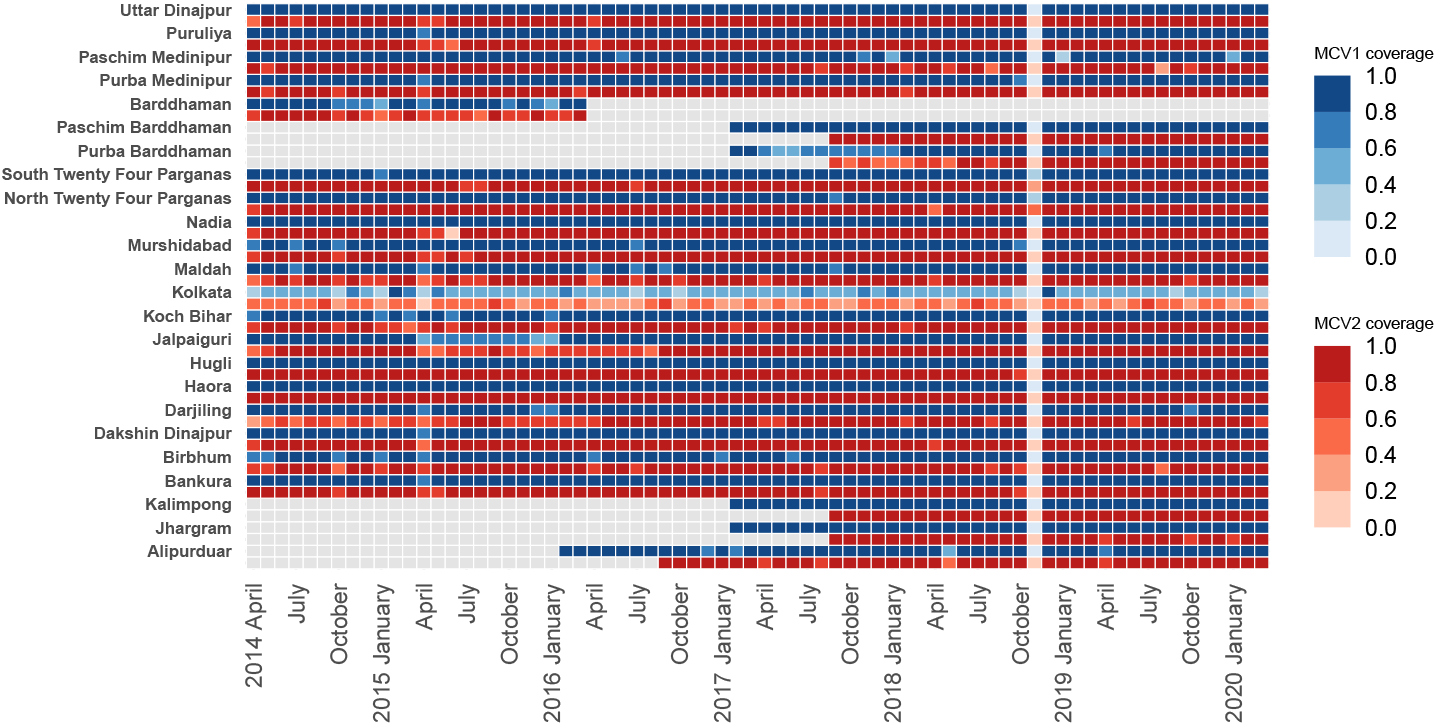
District wise vaccination coverage in West Bengal from 2014 to 2020 corresponding to Section 5. Grey shading indicates periods prior to the initiation of vaccination or missing vaccination data for specific districts. Imputation was used for the missing coverages,, corresponding to Section 5.

As the first dose of the measles vaccine is administered at 9 months of age and the second dose at 16 months of age, to account for the vaccination effect, we again took the unvaccinated cohort at time *t* in district *j* as:

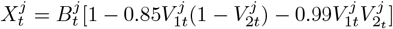

where 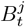 denotes the birth cohort in the time period *t* in district *j*. The respective vaccine coverages, 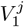 and 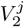 are taken from Figure 5. As discussed in Section 2.3, the generative equation for the number of susceptibles at time *t* is updated to 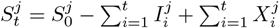.

For setting up the spatial neighborhoods, both choices N1 and N2 were used. In the nearest neighbor method (N2) *k* was taken to be 3. As before, we tested for trend and seasonality terms in our model. We have applied the model described in section 2.1, with three nearest neighbor districts as a spatial cluster (*N*_2_ choice). For model training, data from April 2014 to September 2019 were used, while forecasting was done for October 2019 to March 2020. The prediction model for each district was chosen as described based on several GOFs as described in Section 4. The BIC value was reported to be lowest for *p* = 1 (4329.40). We do not report the parameter estimation results for the 23 districts due to space constraints. In Figures 6a, we show the medium-term forecast of six months in terms of RMSE of few districts covering the entire span of the state level map of West Bengal, with respect to two types of spatial neighbor choices, N1 and N2. For *N*_1_, we used 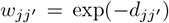. From Figure 6a, we note that the forecast results based on the prediction model with choice N1 outperformed the model that only involved 3 nearest districts (*N*_2_). The same pattern is observed in all districts. This pattern (N1 performs better) is likely to be observed due to the well-developed connectivity between the districts of West Bengal. The movement of individuals may not be restricted to the immediate neighboring units and may extend across all districts. Noting that West Bengal’s 3,825-kilometer rail network, mainly run by the Eastern and South Eastern Railway zones, connects its districts efficiently. Important routes like the Howrah–Barddhaman–Asansol and Malda–New Jalpaiguri lines handle key east-west and north-south travel. The widespread Kolkata Suburban Railway serves as a vital daily link for eight surrounding districts. This rail network is complemented by a robust road system, with major National Highways such as NH-34, NH-2, NH-31, and NH-55 ensuring strong connectivity throughout the state. We also note the MAE and coverage values of the forecasts for the N1 choice in Figure 6b.

**Figure 6:**
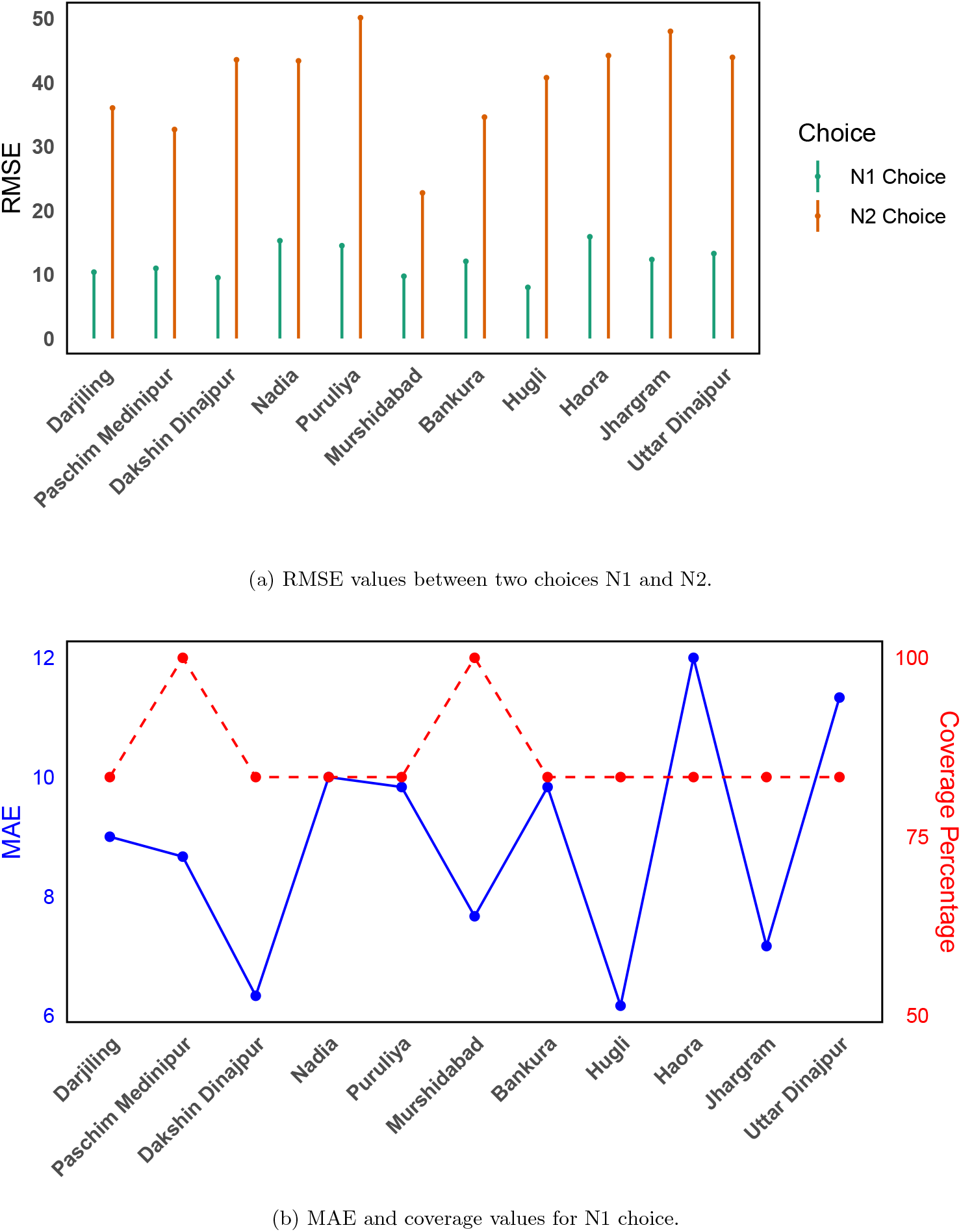
RMSE, MAE and coverage values for few selected districts, spanning the entire state level map of West Bengal, Section 5.

Based on the estimated values of *N*_1_ choice, we sought to identify the dominant inter-district connections influencing measles transmission. Since the choice of *N*_1_ assumes a fully connected network in which all districts are linked, we introduced sparsity by applying a thresholding procedure to the estimated interaction parameters 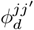. The estimated values of 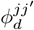, for all district pairs *j, j*^*′*^, ranged from 0 to 0.237, with a median value of 0.014. We classified a connection as dominant if 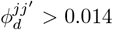; otherwise, the corresponding parameter was set to zero. This thresholding approach enabled the identification of key transmission pathways while reducing weaker connections.

The number of dominant edges associated with each district is illustrated in Figure 7. The districts in the central region of West Bengal exhibit greater connectivity, likely reflecting stronger transportation networks and greater livelihood opportunities that facilitate population movement. In contrast, northern districts show comparatively fewer dominant connections, consistent with migration patterns in which individuals tend to move toward the more economically active central regions of the state.

**Figure 7:**
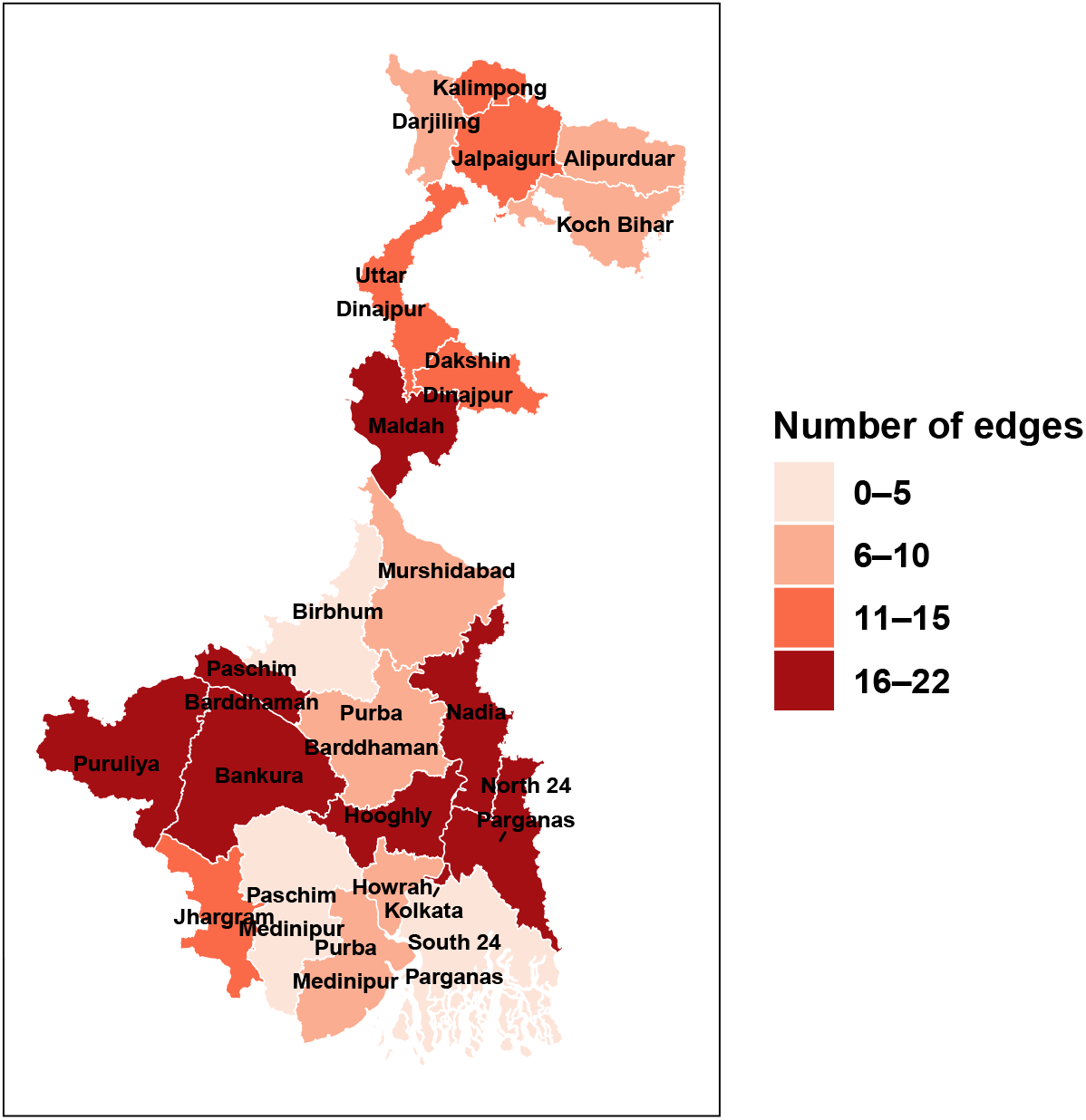
Connection strength of each district in West Bengal corresponding to Section 5.

## 6 Concluding Remarks

In this study, we developed a multi-region chain binomial model to predict and forecast disease counts based on surveillance data. Our approach integrates time series techniques, specifically the generalized linear auto regressive model, within an epidemiological framework. The model selection process involved evaluates multiple candidate models to identify the one that performed the best. Through the simulated and real examples, we were able to show that our proposed model is able to predict and forecast disease counts over various time horizons and also remain robust over model misspecifications. Our model performed comparably to the HHH model when the susceptible population was large, but outperformed it as the susceptible population decreased. We observed an AR(1) model was chosen for both real data sets based on measles counts. This happens as a directly transmitted disease like measles has a welldefined biweekly generation interval which allows the AR(1) model to capture the transmission dynamics along with the susceptible depletion. However, our proposed framework with the AR(p) linear predictor allows a flexible setup for diseases other than measles with delayed transmission effects. We have explored this in the simulated data examples.

In this manuscript, we considered a maximum of 23 spatial regions, corresponding to the West Bengal data set. For larger spatial systems with many neighbouring regions, parameter estimation may become challenging. One possible extension is to incorporate a lasso-type penalty, as proposed by Sathish et al. [2025], to investigate sparse spatial dependence. Another limitation of the current work is that it does not account for under-reporting, a common feature of routine surveillance data. As a future work, we are developing an extension that integrates sero-surveillance data to better estimate the infections. We also plan to combine this approach with penalized estimation to analyse larger spatial networks, such as the 60-city UK data set.

Finally, infectious disease incidence often shows substantial age-specific heterogeneity. This phenomenon is particularly pronounced in childhood infections, where younger children tend to be more vulnerable to infection than older children. In the future, we aim to incorporate such age-specific heterogeneity into our spatio-temporal modelling framework and investigate its impact on parameter estimation as well as short- and long-term forecasting performance.

## Data Availability

All data produced in the present study are available upon request to the authors

https://github.com/pallabksinha/Multivariate_chain_binomial_modelling.git

https://hmis.mohfw.gov.in/\#/standardReports

https://github.com/msylau/measles_competing_risks/tree/master/data/raw_data

## 7 Data Sets and Computer Programs

The UK cities measles data set can be downloaded from UK 60 cities data set, while the West Bengal measles data set is publicly available through the Health Management Information System (HMIS), Ministry of Health and Family Welfare, Government of India, and can be downloaded from https://hmis.mohfw.gov.in/#/standardReports. The R program implementing the proposed methodology for case 1 of the simulation study outlined in Section 3.1 is available at Github Link for Codes.

## Disclaimer

The work/opinion is based on research findings by the authors and not the opinion of any government.

## Acknowledgments

We thank Dr. Shubham Niphadkar for his support. Funding for this study was provided by the Gates Foundation (INV-044445). We thank the two anonymous referees for their valuable inputs.

